# Predicting age from 100,000 one week-long 100Hz wrist accelerometer records of physical activity

**DOI:** 10.1101/2021.06.21.21259265

**Authors:** Alan Le Goallec, Sasha Collin, M’Hamed Jabri, Samuel Diai, Théo Vincent, Chirag J. Patel

## Abstract

Physical activity improves quality of life, physical health and mental health, and is also an important protective factor against highly prevalent age-related diseases such as cardiovascular diseases, diabetes, cancer and mental health. With age, physical activity tends to decrease, leading down a vicious cycle that increases vulnerability to disease in the elderly. In the following, we trained neural network architectures to predict age from 115,456 one week-long 100Hz wrist accelerometer recordings from the UK Biobank (R-Squared=63.5±2.4%; root mean squared error=4.7±0.1 years). We achieved this performance by preprocessing the raw data as 2,271 scalar features, 113 time series and four images. We also considered the raw signal at different time scales (weekly activity patterns vs. gait). We then defined accelerated aging for a participant as being predicted older than one’s actual age and aimed to characterize these participants. We performed a genome wide association on the accelerated aging phenotypes to estimate its heritability (h_g^2^=12.3±0.9%) and identified nine single nucleotide polymorphisms in seven genes associated with it (e.g HIST1H4L, involved in chromatin organization). Similarly, we identified biomarkers (e.g blood pressure), clinical phenotypes (e.g chest pain), diseases (e.g hypertension), environmental (e.g smoking) and socioeconomic (e.g income and education) variables associated with accelerated aging. We conclude that physical activity-derived biological age is a complex phenotype with both genetic and non-genetic factors.

## Introduction

Physical activity improves quality of life, physical health and mental health, and is also an important protective factor against highly prevalent age-related diseases such as cardiovascular diseases, diabetes, cancer and mental health ^1^ and reduces risk for mortality ^2^. Despite this crucial importance, physical activity tends to decrease by 40-80% with age ^3^, making the elderly more vulnerable to the aforementioned diseases.

In the era of big data, new sensors such as accelerometers can be used to capture the level of physical activity of individuals throughout the day, providing researchers with new opportunities to understand the link between physical activity and healthy aging. Two publications for example leveraged the NHANES dataset to predict age ^4,5^. As gait changes as one gets older ^6^, several research teams used this feature to predict chronological age ^7–15^. There have been efforts to predict components of “physical fitness”, such as cardiorespiratory health in mortality ^16^. Finally, use of newer tools, such as accelerometers, have been proposed: Strain et. al built a survival model on UKB’s wrist accelerometer data ^17^ predicting all-cause mortality. We are, to our knowledge, the first to use data from participants of UKB to predict age.

In the following, we leveraged 115,456 one week-long wrist accelerometer records collected from 103,680 participants aged 45-81 years to shed light on the complex relationship between aging and physical activity. We preprocessed the raw data into scalars, time series, and “images” (Figure 1A, B, C and D), that we then used as predictors to train deep neural network architectures to predict age. We defined the biological age of a participant as the prediction outputted by the model.

**Figure 1:**
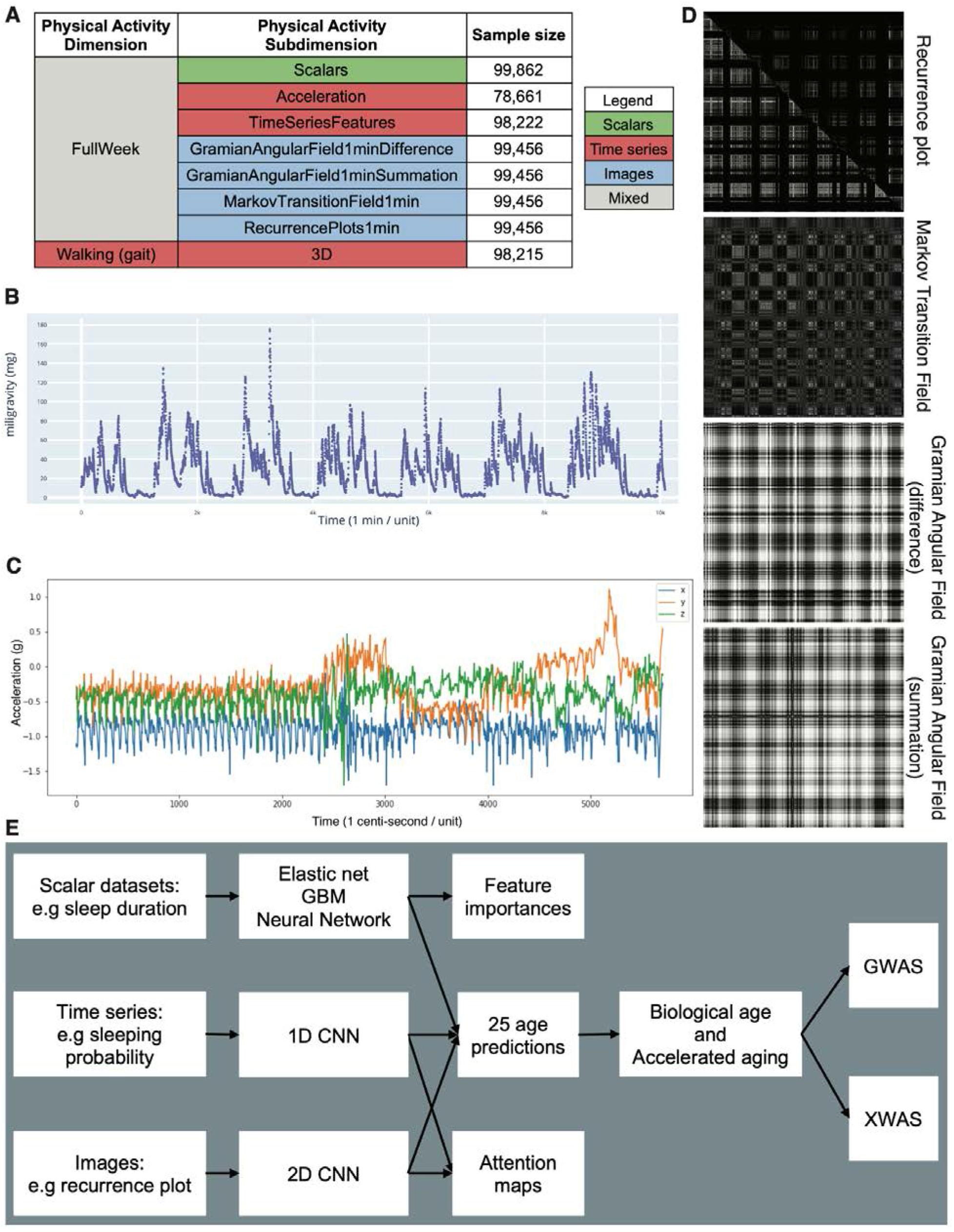
Presentation of the datasets and overview of the analytic pipeline. **A - Preprocessed datasets obtained from the raw wrist accelerometer data, hierarchically ordered into physical activity dimensions and subdimensions. B - full week - Acceleration sample. Each time step corresponds to the mean acceleration over one minute. C - Gait sample, collected over one minute. D - Images generated from a raw accelerometer record. E - Overview of analytic pipeline.** D - See methods for a description of the images. For example, the recurrence plot measures the similarity between the physical activity captured at different times of the week. The axes each correspond to the full week. The upper right triangle corresponds to similarities in sleep activities, and the lower left triangle corresponds to similarities in the remaining time.

In contrast to chronological age --the time since an individual’s birth--, biological age represents the state and condition of the individual’s body and is the true underlying cause of age-related diseases. We defined accelerated aging as being predicted older than one’s age. Second, we performed a genome wide association study [GWAS] to estimate the heritability of the phenotype, as well as to identify single nucleotide polymorphisms [SNPs] associated with the new phenotype . Similarly, we performed an X-wide association study [XWAS] to identify biomarkers, clinical phenotypes, diseases, family history, environmental and socioeconomic variables associated with accelerated aging. (Figure 1E)

## Results

### We predicted chronological age within five years

We performed our analysis on the 115,456 one week-long wrist accelerometer records collected from 103,680 UK Biobank participants aged 45-81 years (Fig. S1). First, we preprocessed the raw one week-long 100Hz time series (60,480,000 time steps) into 2,271 scalar variables (e.g average acceleration, number of hours sleeping, number of hours exercising, Table S1), 113 time series with one time step per minute (e.g probability of being asleep, maximum physical activity, minimum physical activity) and four images (recurrence plot, Markov transition field and Gramian angular field: difference and summation). For each sample, we also extracted a one minute-long 100Hz sample for which the participant is walking (Figure 1C). This dataset, referred to as “Walking” (Figure 1A) allows us to test whether age can be predicted from the gait, in contrast to the other datasets who summarize the activity pattern over the full week.

We predicted age from scalar variables using an elastic net, a gradient boosted machine [GBM] and a shallow, fully connected neural network. We predicted age from time series using one-dimensional convolutional neural networks [CNNs] and from images using two-dimensional CNNs. We hierarchically ensembled the models and predicted age with a R-Squared [R^2^] of 63.5±2.4% and a root mean squared error [RMSE] of 4.7±0.1 years. In terms of the different physical activity “dimensions” (full week activities vs. gait), we predicted age with respective R^2^ values of 59.4±4.5% and 39.7±1.3%. Accelerated aging, as defined by the full week activity pattern, was .266±.004 correlated with accelerated aging, as defined by the participant’s gait (Fig. S4). For the models built on the full week data, the best performance was obtained from the GBM trained on the scalar features (R^2^=59.4±0.6%).

We report the feature importances for the elastic net, the GBM and the neural network in Table S2. We generated attention maps for the time series (Fig. S3) and the images (Fig. S2). More examples can be respectively be found at https://www.multidimensionality-of-aging.net/model_interpretability/time_series and https://www.multidimensionality-of-aging.net/model_interpretability/images under “Physical Activity”.

### Genetic factors and heritability of accelerated physical activity-derived aging

We defined the biological age of each participant as their predicted age by the model, and accelerated aging as the difference between their biological age and their chronological age. We then performed a genome wide association study [GWAS] and found accelerated aging to be 12.3±0.9% GWAS-heritable. We identified nine SNPs in seven genes associated with the phenotype. The GWAS highlighted two peaks: (1) RSL24D1P1 (a pseudogene). RSL24D1P1 is in linkage disequilibrium with HIST1H4L (H4 Clustered Histone 13, an histone, involved in chromatin organization); and (2) OR5V1 (Olfactory Receptor Family 5 Subfamily V Member 1, involved in olfactory transduction).

### Non-genetic factors associated with accelerated aging

We performed an X-wide association study [XWAS] to identify biomarkers (Table S3), clinical phenotypes (Table S6), diseases (Table S9), environmental (Table S12) and socioeconomic (Table S15) variables associated with accelerated aging. We found no association between accelerated aging and family history variables. We summarize our findings below. The full results can be exhaustively explored at https://www.multidimensionality-of-aging.net/xwas/univariate_associations.

#### Biomarkers associated with accelerated aging

The three biomarker categories most associated with accelerated aging are blood pressure, anthropometry and pulse wave analysis (a measure of arterial stiffness) biomarkers (Table S4). Specifically, 100.0% of blood pressure biomarkers are associated with accelerated aging, with the three largest associations being with pulse rate (correlation=.082), systolic blood pressure (correlation=.041), and diastolic blood pressure (correlation=.024). 62.5% of anthropometry biomarkers are associated with accelerated aging, with the three largest associations being with waist circumference (correlation=.114), hip circumference (correlation=.094), and weight (correlation=.092). 50.0% of arterial stiffness biomarkers are associated with accelerated aging, with the three largest associations being with pulse rate (correlation=.078), position of the pulse wave notch (correlation=.063), and position of the shoulder on the pulse waveform (correlation=.046).

Conversely, the three biomarker categories most associated with decelerated aging are symbol digit substitution (a cognitive test), hand grip strength and spirometry (Table S5). Specifically, 100.0% of symbol digit substitution biomarkers are associated with decelerated aging, with the two associations being with the number of symbol digit matches attempted (correlation=.096) and the number of symbol digit matches made correctly (correlation=.088). 100.0% of hand grip strength biomarkers are associated with decelerated aging, with the two associations being with right hand grip strength (correlation=.088) and left hand grip strength (correlation=.086). 100.0% of spirometry biomarkers are associated with decelerated aging, with the three associations being with forced expiratory volume in one second (correlation=.074), forced vital capacity (correlation=.071), and peak expiratory flow (correlation=.057).

#### Clinical phenotypes associated with accelerated aging

The three clinical phenotype categories most associated with accelerated aging are chest pain, breathing and claudication (Table S7). Specifically, 100.0% of chest pain clinical phenotypes are associated with accelerated aging, with the three largest associations being with chest pain or discomfort walking normally (correlation=.045), chest pain or discomfort (correlation=.043), and chest pain due to walking ceases when standing still (correlation=.042). 100.0% of breathing clinical phenotypes are associated with accelerated aging, with the two associations being with shortness of breath walking on level ground (correlation=.087) and wheeze or whistling in the chest in the last year (correlation=.032). 53.8% of claudication clinical phenotypes are associated with accelerated aging, with the three largest associations being with leg pain on walking (correlation=.069), leg pain when walking uphill or hurrying (correlation=.069), and leg pain when standing still or sitting (correlation=.068).

Conversely, the three clinical phenotype categories most associated with decelerated aging are cancer screening (most recent bowel cancer screening: correlation=.028), mouth health (no mouth or teeth dental problem: correlation=.023) and general health (no weight change during the last year: correlation=.023). (Table S8)

#### Diseases associated with accelerated aging

The three disease categories most associated with accelerated aging are cardiovascular, a category encompassing various diseases (category R, see Table S9) and gastrointestinal diseases (Table S10). Specifically, 40.3% of cardiovascular diseases are associated with accelerated aging, with the three largest associations being with hypertension (correlation=.098), chronic ischaemic heart disease (correlation=.051), and atrial fibrillation and flutter (correlation=.046). 39.8% of the various diseases category are associated with accelerated aging, with the three largest associations being with pain in the throat and chest (correlation=.041), nausea and vomiting (correlation=.037), and abdominal and pelvic pain (correlation=.036). 34.7% of gastrointestinal diseases are associated with accelerated aging, with the three largest associations being with gastro-intestinal reflux disease (correlation=.041), diaphragmatic hernia (correlation=.037), and cholelithiasis (correlation=.032).

We found no disease significantly associated with decelerated aging.

#### Environmental variables associated with accelerated aging

The three environmental variable categories most associated with accelerated aging are sleep, sun exposure and smoking (Table S13). Specifically, 57.1% of sleep variables are associated with accelerated aging, with the three largest associations being with napping during the day (correlation=.095), sleeplessness/insomnia (correlation=.063), and daytime dozing/sleeping-narcolepsy (correlation=.058). 30.0% of sun exposure variables are associated with accelerated aging, with the three largest associations being with looking approximately one’s age (correlation=.055), not knowing whether we look younger or older in terms of facial aging (correlation=.046), and time spent outdoors in the summer (correlation=.043). 29.2% of smoking variables are associated with accelerated aging, with the three largest associations being with number of cigarettes currently smoked daily (correlation=.054), currently smoking tobacco on most or all days (correlation=.054), and difficulty not smoking for one day (correlation=.053).

Conversely, the three environmental variable categories most associated with decelerated aging are electronic devices, physical activity (questionnaire) and smoking (Table S14). Specifically, 40.0% of electronic devices variables are associated with decelerated aging, with the three largest associations being with weekly usage of mobile phone in the last three months (correlation=.089), length of mobile phone use (correlation=.073), and hands-free device/speakerphone use with mobile phone in the last three months (correlation=.064). 37.1% of physical activity questionnaire variables are associated with decelerated aging, with the three largest associations being with usual walking pace (correlation=.141), types of physical in the last four weeks: strenuous sports (correlation=.113), and frequency of strenuous sports in the last four weeks (correlation=.102). 25.0% of smoking variables are associated with decelerated aging, with the three largest associations being with time from waking to first cigarette (correlation=.057), age started smoking in current smokers (correlation=.054), and not currently smoking tobacco (correlation=.045).

#### Socioeconomic variables associated with accelerated aging

The three socioeconomic variable categories most associated with accelerated aging are sociodemographics, social support and household (Table S16). Specifically, 28.6% of sociodemographic variables are associated with accelerated aging, with the two associations being with private healthcare (correlation=.043) and receiving attendance allowance (correlation=.033). 25.0% of social support variables are associated with accelerated aging, with the two associations being with leisure/social activities: religious group (correlation=.084) and not having social activities (correlation=.040). 15.0% of household variables are associated with accelerated aging, with the three largest associations being with owning one’s accommodation (correlation=.091), gas or solid fuel cooking/heating: a gas fire that you use regularly in winter time (correlation=.043), and renting one’s accommodation from local authority, local council or housing association (correlation=.043).

Conversely, the three socioeconomic variable categories most associated with decelerated aging are employment, social support and household (Table S17). Specifically, 38.1% of employment variables are associated with decelerated aging, with the three largest associations being with length of working week for main job (correlation=.238), current employment status: in paid employment or self-employed (correlation=.210), and frequency of traveling from home to job workplace (correlation=.204). 25.0% of social support variables are associated with decelerated aging, with the two associations being with attending sports club or gym (correlation=.087) and attending a pub or social club (correlation=.049). 15.0% of household variables are associated with decelerated aging, with the three largest associations being with total household income before tax (correlation=.142), owning one’s accommodation with a mortgage (correlation=.115), and number of vehicles in household (correlation=.063).

## Discussion

We predicted chronological age from wrist accelerometer records (R^2^=63.5±2.4%; RMSE=4.7±0.1 years). Specifically, we predicted age from both the full week activity pattern (R^2^=59.4±4.5%) and the gait (R^2^=39.7±1.3%) of the participants, and found the two accelerated aging phenotypes derived from these models to be mildly correlated (.266±.004). This suggests that both weekly physical activity patterns and gait are changing with age, but that a single individual might for example have a gait more typical of someone younger than them, despite having a weekly physical activity pattern more typical of someone older than them.

Interestingly, despite the large sample size (N=98,222) and contrary to our experience, the gradient boosted machine model trained on the scalar predictors outperformed the convolutional neural network trained on the time series (R^2^=59.4±0.6% vs. 41.8±0.6%).

We performed a GWAS and found that accelerated aging is partly heritable (h_g^2^=12.3±0.9%) with nine SNPs in seven genes associated with the phenotype, with tentative signals in chromosome six located close and or within the human leukocyte antigen (HLA) coding region, known to play a critical role in variation in immune response and autoimmune disease risk ^18^.

Accelerated aging is associated with biomarkers, clinical phenotypes linked to cardiovascular (blood pressure, arterial stiffness, electrocardiogram, pulse wave analysis, chest pain, cardiovascular diseases), pulmonary (spirometry, breathing, blood count, pulmonary diseases, anaemia), musculoskeletal (hand grip strength, heel bone densitometry, claudication, joint pain, arthritis, falls, fractures) and metabolic health (anthropometry, impedance, blood biochemistry, diabetes, obesity and other metabolic diseases). These associations with cardio-respiratory ^19,20^, metabolic ^21^ and musculoskeletal health ^22^ confirm the biological relevance of accelerated aging as measured by physical activity.

Additionally, accelerated aging is associated with biomarkers, clinical phenotypes and diseases in physiological dimensions that are not as directly related to physical activity, such as cognitive function, brain MRI features, mental health, brain diseases, hearing, eyesight, eye diseases, gastro-intestinal diseases. These associations could be explained by their indirect effect of the physical activity pattern of the participant. Another explanation is that accelerated cardiovascular, musculoskeletal and metabolic aging is part of a general aging process, and that accelerated aging as captured by an accelerometer is correlated with accelerated aging in all organs, even the ones that are not directly involved in physical activity. This second hypothesis is supported by the association between wrist accelerometer-derived accelerated aging and general health factors (e.g general health rating, long-standing illness disability or infirmity, personal history of medical treatment or disease). We explored the connection between wrist accelerometer-derived aging and aging of other organ systems in a different paper ^23^.

In terms of environmental exposures, accelerated aging is associated with self-reported sleep, time spent outdoors, time spent on electronic devices and physical activity. These associations are expected since these features were leveraged by the model to predict chronological age in the first place. Accelerated aging is also associated with smoking, diet (e.g processed meat intake), alcohol intake (e.g frequency) and medication (e.g taking a cholesterol-lowering medication), which have been linked to cardiovascular health ^24^. We also found that being breastfed as a baby and being comparatively shorter at age ten is associated with decelerated aging, with the latter association being potentially explained by the fact that during youth and early adulthood, taller height is associated with higher blood pressure. In the literature, height is reported to be negatively correlated with blood pressure ^25^, but in the UK Biobank cohort, the correlation is positive until approximately sixty-five years ^26^. Interestingly, decelerated aging is also associated with looking younger than one’s age (correlation=.084).

Socioeconomic status (sociodemographics, social support, household, education, employment) is negatively associated with accelerated aging, reflecting the literature. For example, the richest 1% live significantly longer than the poorest 1% in the US (males: 14.6±0.2 years longer; females: 10.1±0.2 years longer) ^27^.The association is likely mediated in part by differences in access to health care and health literacy ^28^.

The XWAS identified candidate lifestyle interventions to slow aging (e.g for diet: salad/raw vegetable intake, eating butter as opposed to margarine), but an important limitation of our study is that UKB is an observational study and that correlation does not necessarily imply causation. A future direction could be to leverage participants with repeated measurements to test whether a change in lifestyle (e.g change of diet) between the two measurements led to slower biological aging.

We compare our physical activity-derived age predictors with the ones already existing in the literature in the supplementary. A summary of this comparison can be found in Table S19.

Our predictor could be used to analyze the data collected from smartwatches and fitness trackers to provide users with an estimation of their biological age and identify individuals at risk for diseases (e.g cardiovascular, pulmonary, musculoskeletal and metabolic). Our predictor could also be adapted to perform similar tasks from smartphone accelerometer data. Finally, a new horizon includes testing whether emerging rejuvenating therapies ^29^ would be associated with a reduction of biological age as defined by our wrist accelerometer-based model.

## Methods

### Data and materials availability

We used the UK Biobank (project ID: 52887). The code can be found at https://github.com/Deep-Learning-and-Aging. The results can be interactively and extensively explored at https://www.multidimensionality-of-aging.net/. We will make the biological age phenotypes available through UK Biobank upon publication. The GWAS results can be found at https://www.dropbox.com/s/59e9ojl3wu8qie9/Multidimensionality_of_aging-GWAS_results.zip?dl=0.

### Software

Our code can be found at https://github.com/Deep-Learning-and-Aging. For the genetics analysis, we used the BOLT-LMM ^30,31^ and BOLT-REML ^32^ software. We coded the parallel submission of the jobs in Bash ^33^.

### Cohort Dataset: Participants of the UK Biobank

We leveraged the UK Biobank ^34^ cohort (project ID: 52887). The UKB cohort consists of data originating from a large biobank collected from 502,211 de-identified participants in the United Kingdom that were aged between 37 years and 74 years at enrollment (starting in 2006). 103,688 UKB participants received wrist accelerometers that they wore for a week to measure their physical activity levels^35^. Some of them repeated the experience during up to four “seasonal repeats” to evaluate the variability of their physical activity pattern between seasons, so the total number of accelerometer records amounts to 115,464. The raw data is available and consists of three leads (x, y and z axes) and a sampling frequency of 100 Hz with a dynamic range of ±8g (field 90001). A preprocessed version of the data is also available and consists of one lead that reports the average acceleration every five seconds, after filtering for noise and correcting for gravity (field 90004). For the preprocessed version, only one sample per participant is available, for a total of 103,687 samples.

The Harvard internal review board (IRB) deemed the research as non-human subjects research (IRB: IRB16-2145).

### Definition of the different physical activity dimensions

We defined physical activity dimensions by hierarchically grouping the different datasets at two different levels: we call the first level “physical activity dimensions” and the second level “physical activity subdimensions”. The complete hierarchy between the two physical activity dimensions and the eight subdimensions is described in Figure 1A . For example, one of the physical activity dimensions we investigated was “Full Week”, which is based on the participants’ physical activity over the entire week. This dimension consists of seven subdimensions: (1) the scalar features derived from the full week accelerometer data (e.g mean acceleration over the week, maximum acceleration during the weekend), (2-3) the models built on the full week time series generated from the raw data, and (4-7) the images generated from the raw data.

### Data types and Preprocessing

The data preprocessing step is different for the different data modalities: demographic variables, scalar predictors, time series and images. We define scalar predictors as predictors whose information can be encoded in a single number, such as the mean acceleration, as opposed to data with a higher number of dimensions such as time series (one dimension, which is time), and images (two dimensions, which are the height and the width of the image).

#### Demographic variables

First, we removed out the UKB samples for which age or sex was missing. For sex, we used the genetic sex when available, and the self-reported sex when genetic sex was not available. We computed age as the difference between the date when the patient attended the assessment center and the year and month of birth of the patient to estimate the patient’s age with greater precision. We one-hot encoded ethnicity.

#### Physical activity as time series

First, we preprocessed the data to ensure we were analyzing activity that took place during the full week. To mitigate the problems associated with excessively long time series, we started with the summarized data (one lead, 0.2 Hz, field 90004) and computed the mean acceleration every minute. We discarded the samples for which less than a full week’s worth of data was available (10,080 time steps = 7 days * 24 hours * 60 minutes), and only kept the first 10080 when more data than necessary was available. This step filtered out 25,026 samples out of the initial 103,687 samples, leaving 78,661 samples. Finally, we applied a smoothing function (exponential moving average with parameter 35) to reduce the noise. We refer to this preprocessed data as “FullWeek_Acceleration” (Fig. S5).

Then, we generated a second time series (“FullWeek_TimeSeriesFeatures”) over the full week by processing the raw high frequency 3D data (field 90001). We used the “biobank Accelerometer Analysis” software ^36,37^ to compute the probability for each sample of performing several activities (biking, sitting/standing, vehicle, mixed, sleeping and walking) every five seconds. The software generates 113 features as an intermediate step, that it uses in a later step to classify the different activities. We computed the average for these time series every 5 seconds to leverage these features as additional channels for the FullWeek_TimeSeriesFeatures model. The final dimensions of each sample was therefore 113 channels of 7 days * 24 hours * 12 5-minutes long periods = 5,373 time steps.

We then filtered low quality samples. First, we removed 6,618 samples for which data was collected for less than a week. The summary file outputted by the software that we used to preprocess the data also contains quality indicators. We used four columns (quality-calibratedOnOwnData, quality-goodCalibration, quality-goodWearTime and quality-daylightSavingsCrossover) to filter out 8,984 low quality samples. After preprocessing the dataset, we were left with 99,862 samples.

We generated a third time series from the raw high frequency 3D data to specifically compare the walking pattern between the different participants, which we refer to as “Walking_3D”. We identified for each sample the continuous periods during which the activity was predicted by the software to be walking with a probability of 1. For each sample, we then randomly selected up to five of these sequences and extracted the corresponding 3D raw acceleration data (Fig. S6).

Finally, we generated a similar dataset to predict age as a function of physical activity during sleeping. The predictor was weak so it is not included in the main tables, but we describe our methods for researchers who would be interested in reusing this pipeline for different purposes. First we detected continuous periods of 180 and 360 minutes of sleep. Because the probability of the sleeping activity is never equal to 1 over these long periods of time, we first generated binary time series that specified every five seconds whether the activity was sleeping with a probability of 1, or not. Then we smoothed this curve by using a sliding window with a period of 30 minutes and set a threshold of 0.3 to determine the beginning and the end of the sleeping period. This allowed us to include brief periods of physical activity during the night as being part of the sleeping period. We then extracted the 3D high intensity acceleration data corresponding to the sleeping periods of 180 and 360 minutes. We extracted as many of these samples as possible for each participant. Neither the 180 minutes nor the 360 minutes hyperparameters yielded good predictors of chronological age (R-Squared=0.016 for both models).

#### Physical activity as scalar features

We define scalar data as a variable that is encoded as a single number, such as mean or maximum acceleration, as opposed to data with a higher number of dimensions, such as time series, and images. The complete list of scalar biomarkers can be found in Table S2. We did not preprocess the scalar data, aside from the normalization that is described under cross-validation further below.

We filtered out the low-quality samples based on the quality-calibratedOnOwnData, quality-goodCalibration, quality-goodWearTime and quality-daylightSavingsCrossover indicators, as mentioned in “Physical activity as time series” above. Then we generated a total of 2,271 scalar features, from three different pipelines (Table S1).

First, we leveraged 1,333 summary features directly outputted by the two biobank Accelerometer Analysis preprocessing softwares ^36,37^. The first software ^37^ generated 813 features from which we filtered out 66 (Table S18) non-biological features (e.g. features related to the calibration of the accelerometers) and kept the remaining 747 features. The second software ^36^ generated 896 features, from which we filtered out 67 non-biological features. We then took the union of the features generated by the two models to obtain the 1,333 features.

Second, we used the 113 time series generated by the Accelerometer Analysis software during one of its intermediate steps. For each of these time series, we computed the minimum value, the maximum value, the average value, the standard deviation, as well as the 25th, 50th and 75th percentiles, for a total of 791 features.

Finally, we built 147 custom features from six different types. (1) Summary statistics features, (2) Behavioral features, (3) Sleeping features, (4) Activity phase features, (5) Weekend features, (6) Metabolic Equivalent of Task (MET) features. The design of these features is described in detail below.

##### Summary statistics features

First, we computed the average acceleration every minute for each participant, and we computed the mean, median, interquartile range, skewness, and the fifth, tenth, 25th, 75th and 90th percentile on these time series. We then computed the average acceleration over every day of the week, and we computed the following summary statistics: the mean, standard deviation, minimum, maximum, median, and the 25th and 75th percentiles. Finally, we computed the same summary statistics on the different periods of the day: (1) night (midnight to 6am), (2) morning (6am to 9am), (3) day (9am to 7pm) and (4) evening (7pm to midnight).

##### Activity specific features

We computed the number of minutes spent on each activity (e.g. walking, sleeping). We then computed the distribution of time across the different activity types for each day of the week along with the standard deviation for these values. Finally, we computed the same features described under “Summary statistics features” for every physical activity. For each activity, we defined the time series on which to perform this analysis as the concatenation of periods of at least five minutes over which the activity classification remained constant.

##### Sleeping features

We defined night sleep as time steps classified as sleep between 10pm and 6am, and we computed the average night sleep duration. We defined deep sleep as an uninterrupted series of timesteps classified as sleeping, and we computed the average duration of deep sleep phases for each participant. Two participants could have the same average night sleep duration of eight hours, but the first participant could have a single deep sleep phase of eight hours while the second participant could briefly wake up or move in their sleep three times throughout the night, which would read as an average deep sleep duration of only two hours. The average deep sleep duration can therefore be used to assess the quality of sleep that each participant is getting. Then, we defined a sleep session as a sleeping activity longer than twenty minutes and we computed the daily average number of sleep sessions for each participant. This feature for example captures if a participant is taking naps. Finally, we selected the longest sleep session for each participant, which represents their “night sleep”, even if some participants might be working night shifts and therefore have this “night sleep” during the day. We computed the average duration of this night’s sleep and the average time of the day at which it started for each participant.

##### Physical activity features

We computed the daily average time spent walking, as well as the daily average duration of the longest continuous walk. We also computed the average time of the day associated with the first and the last peak of physical activity.

##### Weekend features

We defined the weekend as the time between Friday 10pm and Sunday 11:59pm and we computed the time spent on the different activities during this period. We also computed the time at which the participants went to bed on the night of Saturday evening to Sunday morning to capture Saturday night habits.

##### Metabolic Equivalent of Task (MET) features

The metabolic equivalent of a task is the rate at which a participant is expending energy when performing this task compared to when they are resting (e.g. sitting or being inactive) ^38^. For example, a MET of two means that the participant is spending twice as much energy performing their current activity than they are when they are resting. Activities with a MET smaller than three (e.g. walking slowly or working at a desk) are considered to be light intensity activities. Activities with a MET between three and six qualify as moderate intensity activities (e.g. walking at a faster pace or practicing relaxing sportive activities). Activities with a MET larger than six are considered vigorous intensity activities (e.g. jogging, biking, dancing, weightlifting, competitive sportive activity).^38–42^

MET measures can be used to estimate the total energy expended over the day or the week, using the unit of MET hour. For example, if a participant performs a task with a MET of three for 40 minutes, that is equivalent to performing a task with a MET of one for two hours. In both cases, the total energy expended by the participant while practicing the activity is two MET hours.

The UKB software estimates the MET for every time step when it preprocesses the data. We leveraged this column to compute the following features. First, we computed the total MET hours over the full week. Then, we computed the daily (seven values) and the hourly (24 values) MET averages and associated standard deviations. Finally, we computed the mean and the standard deviation of the MET values for low, medium and vigorous activities over the week. We defined low, medium and vigorous intensity using the aforementioned threshold values.

#### Physical activity as images

Starting from the unidimensional, preprocessed acceleration average with a frequency of 0.2 Hz (field 90004), we computed the average acceleration for every minute. From there, we generated four “images” per sample for each participant. We describe how these “images” are created below.

For the first, we generated a custom recurrence plot^43^. We used a sliding window of length ten minutes and stride of one minute, so the value of the pixel in row i and column j is the Euclidean distance between the vector of the 10 one-minute-average accelerations starting at the minute i and the vector of the 10 one-minute-average accelerations starting at the minute j and stored them as floats. We generated two recurrence plots. One using only the time steps for which the probability of the participant sleeping was 1, and one for the remaining time steps. Because recurrence plots are symmetrical, we merged these two recurrence plots into a single image, in which the upper right triangle corresponds to the sleeping timesteps, and the lower left triangle corresponds to the remaining time steps (Supplementary Figure 67).

Second, we generated a Markov transition fields image (n_bins=8), a Gramian angular difference fields image and a Gramian angular summation field image^44^ respectively using the MarkovTransitionField and the GramianAngularField functions of the pyts python package^45^. Finally, we resized all the images to be of size 316*316. A sample of these three images can be found in Supplementary Figure 67.

We generated other images that we ended up not using for the analysis. We will describe them here as knowing that they were poor age predictors might save time for researchers considering similar approaches. First, we generated the recurrence plots using the average acceleration over periods of 32 minutes instead of one minute, which generated images of dimension 316*316 directly, saving the resizing step. We did not observe significant differences between the two methods when we explored this possibility, so we discarded it. We also applied the binary filter traditionally used for recurrence plots, by setting to one the value for the pairs of points that were in the 95th percentile for proximity. We found that the performances were similar, with a possible advantage for using float values. Finally, we also applied these four methods (recurrence plot, Markov transition field and Gramian angular summation and Gramian angular difference) to selected segments of 3D data for the walking and the sleeping activities, possibly capturing the recurrence patterns in the gait, for example. During our preliminary analysis and early exploration of the hyperparameters, we found that none of these 3D data-based plots yielded R-Squared values higher than 0.05.

### Machine learning algorithms

For scalar datasets, we used elastic nets, gradient boosted machines [GBMs] and fully connected neural networks. For times series, images and videos we used one-dimensional, two-dimensional, and three-dimensional convolutional neural networks, respectively.

#### Scalar data

We used three different algorithms to predict age from scalar data (non-dimensional variables, such as laboratory values). Elastic Nets [EN] (a regularized linear regression that represents a compromise between ridge regularization and LASSO regularization), Gradient Boosted Machines [GBM] (LightGBM implementation ^46^), and Neural Networks [NN]. The choice of these three algorithms represents a compromise between interpretability and performance. Linear regressions and their regularized forms (LASSO ^47^, ridge ^48^, elastic net ^49^) are highly interpretable using the regression coefficients but are poorly suited to leverage non-linear relationships or interactions between the features and therefore tend to underperform compared to the other algorithms. In contrast, neural networks ^50,51^ are complex models, which are designed to capture non-linear relationships and interactions between the variables. However, tools to interpret them are limited ^52^ so they are closer to a “black box”. Tree-based methods such as random forests ^53^, gradient boosted machines ^54^ or XGBoost ^55^ represent a compromise between linear regressions and neural networks in terms of interpretability. They tend to perform similarly to neural networks when limited data is available, and the feature importances can still be used to identify which predictors played an important role in generating the predictions. However, unlike linear regression, feature importances are always non-negative values, so one cannot interpret whether a predictor is associated with older or younger age. We also performed preliminary analyses with other tree-based algorithms, such as random forests ^53^, vanilla gradient boosted machines ^54^ and XGBoost ^55^. We found that they performed similarly to LightGBM, so we only used this last algorithm as a representative for tree-based algorithms in our final calculations.

#### Time series

We developed a different architecture for the three time series extracted from the wrist accelerometers data.

##### Architecture for raw acceleration data over the full week

For the model based on the acceleration data across the full week (FullWeek acceleration), we used the architecture from the paper “Extracting biological age from biomedical data via deep learning: too much of a good thing?” ^4^. The architecture of the model is described in Fig. S8 and Fig. S9 and is constituted of two blocks: (1) a convolutional block and (2) a dense block. These two blocks form a seven layers deep neural network, preceded by a batch normalization. The inputs for the convolutional block are time series with a single channel and 10,080 time steps. The convolutional block consists of four one-dimensional convolutional layers with respectively 64, 32, 32 and 32 filters, a kernel size of respectively 128, 32, eight and eight, and a stride of one. Every convolutional layer is zero-padded and followed by a max pooling with a size of respectively four, four, three and three, and with a stride of two. The output of the convolutional block is inputted into the dense block which consists of three dense layers. The first two dense layers have respectively 256 and 128 nodes and are using dropout. The first six layers of the architecture use kernel and bias regularization, and use ReLU as their activation function. The seventh layer is a dense layer with size one and with a linear activation function to predict chronological age.

During our preliminary analysis, we tested a second architecture, from the article “Deep Learning using Convolutional LSTM estimates Biological Age from Physical Activity” ^5^. This architecture uses both convolutional layers and LSTM layers and takes as input the data formatted as a 3-dimensional data frame of dimensions 7×24×60. The first dimension is for the days of the week, the second for the hours of the day and the third for the minutes of the hour. The model’s architecture can be found in Figure 2 of the aforementioned paper. This architecture explained 33% of the variance in chronological age but was outperformed by the first model described above and took significantly longer to train. As a result, we did not select this second architecture for our final pipeline.

**Figure 2:**
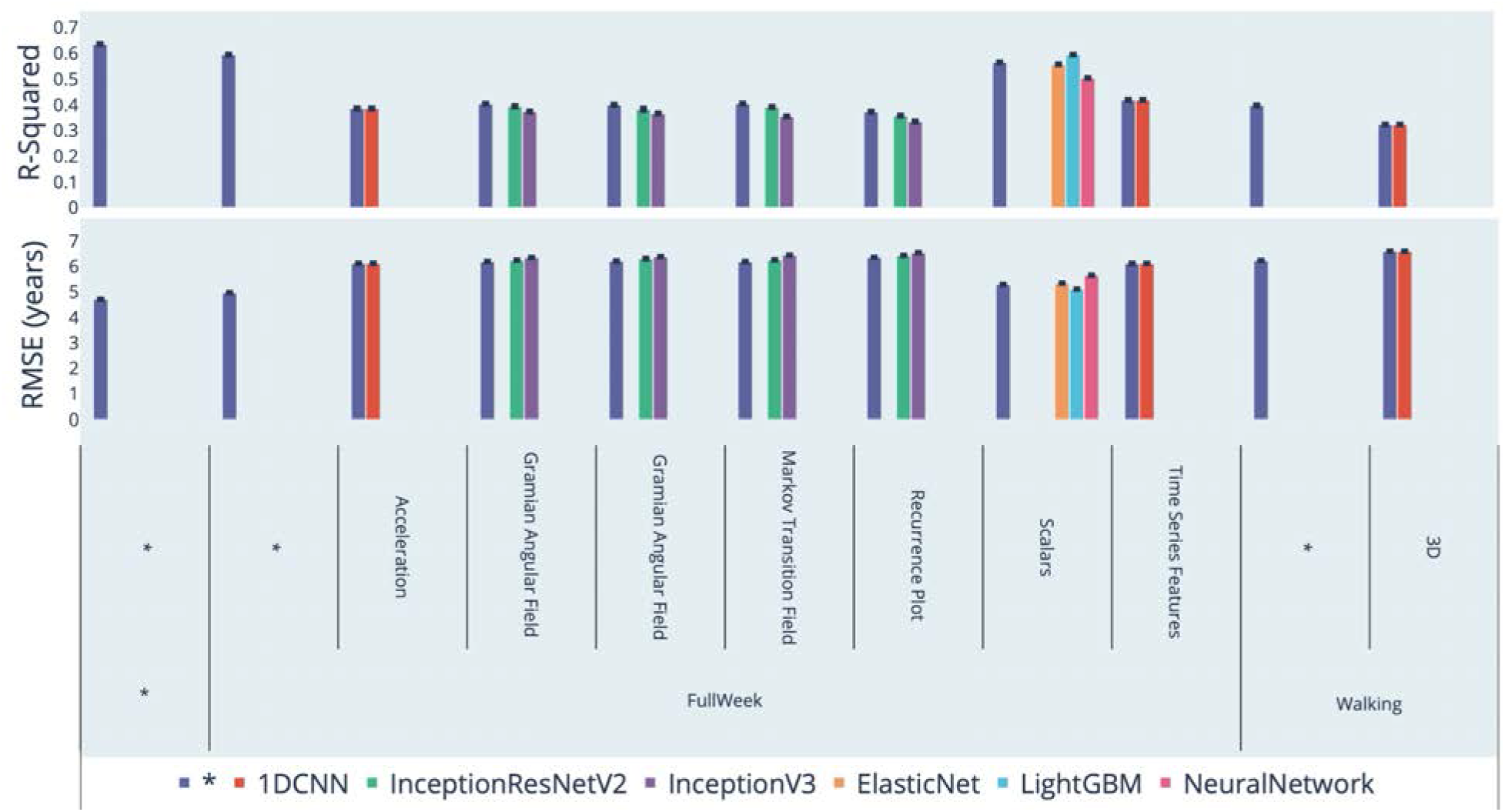
Chronological age prediction performance (R^2^ and RMSE) * represent ensemble models

**Figure 3:**
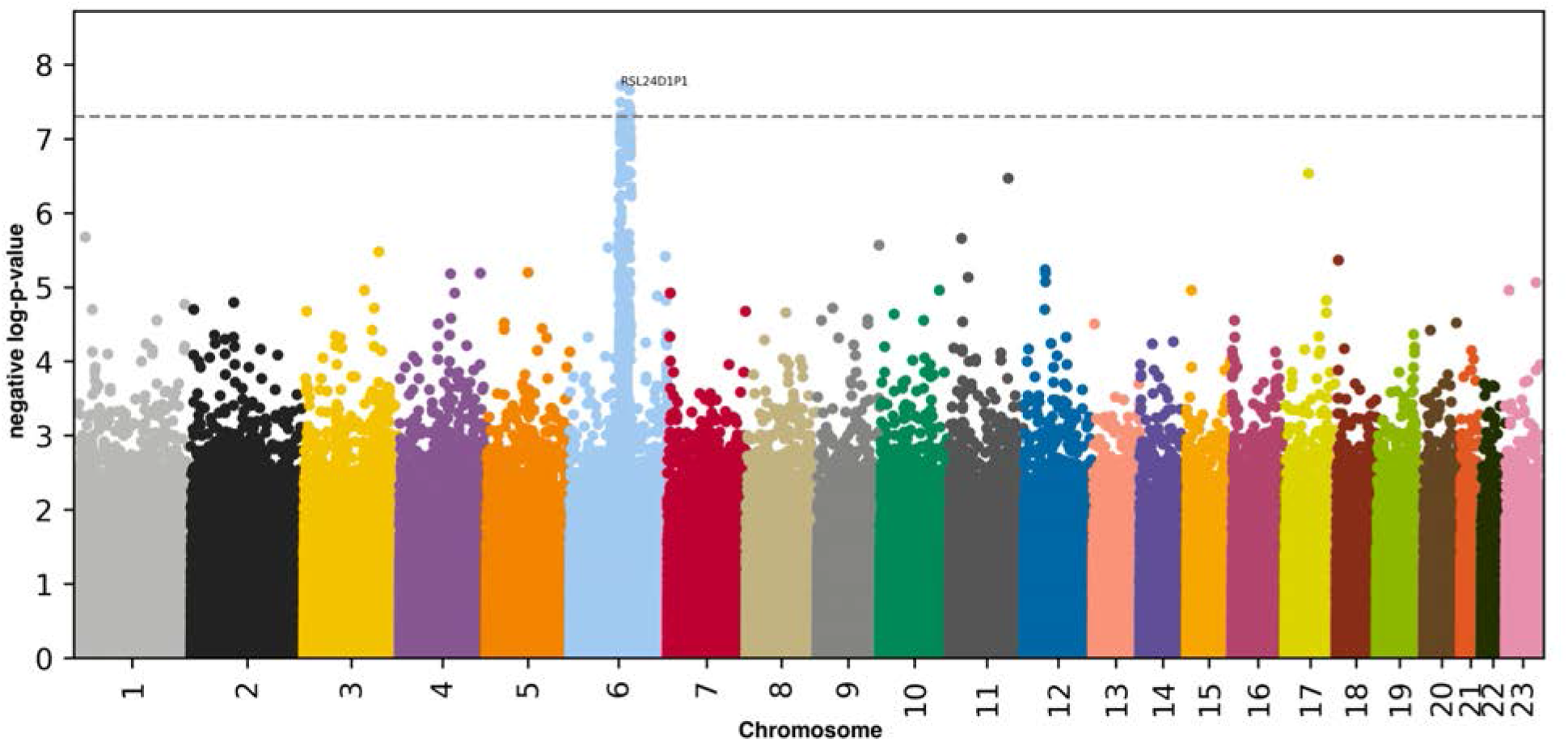
Genome Wide Association Study results. negative log10(p-value) vs. chromosomal position. Dotted line denotes 5e-8.

##### Architecture for features extracted from the acceleration data over the full week

To analyze the 113 time series of features extracted from the raw acceleration data, we built a 14 layers deep architecture which can be found in Fig. S10 and Fig. S11. The architecture consists of a convolutional block and a dense block. The inputs for the convolutional block are time series with 113 channels and 2,016 time steps. The convolutional block comprises nine one-dimensional convolutional layers with the number of filters for each layer doubling with every layer, starting from 64 and with a maximum number of filters of 4,096 (64, 128, 256, 512, 1,024, 2,048, 4,096, 4,096, 4,096). Each convolutional layer is zero-padded and followed by a one-dimensional max pooling layer of size two and stride 2. The output of the convolutional block is converted to 4,096 scalar features using a global max pooling over each of the 4,096 filters. A single layer neural network (409 nodes) is parallelly taking as input the 113 scaling factors for the 113 normalized time series that were inputted in the convolutional block. The 4,096 convolutional based features are concatenated with the 409 scaling factor-based features, and the resulting 4,505 scalar features are inputted in the dense block. The dense block is composed of five dense layers with respectively 1,024, 1,024, 1,024, 512 and one node(s). All dense layers, except for the last one, use dropout. The last layer uses linear activation to predict chronological age. A single layer (25 nodes) neural network is trained on sex and ethnicity, and its output is concatenated with the output of the second dense layer before being inputted in the third dense layer. All the layers of the architecture aside from the final layer use batch normalization, kernel and bias regularization, and ReLU for their activation function.

##### Architecture for three-dimensional high frequency walking data

We built a 13 layers-deep convolutional architecture to analyze the walking data. This architecture can be found in Fig. S12 and Fig. S13. The architecture is highly similar to the one described above. It comprises a convolutional block and a dense block. The inputs for the convolutional block are time series with three channels and 5,700 time steps (5.7 seconds of 3-dimensional acceleration). The convolutional block consists of eight one-dimensional convolutional layers with the number of filters for each layer doubling with every layer, starting from 32 and with a maximum number of filters of 2,048 (32, 64, 128, 256, 512, 1,024, 2,048, 2,048). All convolutional kernels are zero-padded, of size three and with a stride of one. Each convolutional layer uses batch normalization, ReLU as activation function, and is followed by a one-dimensional max pooling layer of size two and stride 2. The output of the convolutional block is converted to 2,048 scalar features using a global max pooling over each of the 4,096 filters. A single layer neural network (204 nodes) is parallelly taking as input the 3 scaling factors for the x-axis, y-axis and z-axis normalized time series that were inputted in the convolutional block (the three dimensions of the accelerometer). The 4,096 convolutional based features are concatenated with the 204 scaling factor-based features, and the resulting 2,252 scalar features are inputted in the dense block. The dense block is composed of five dense layers with respectively 1,024, 1,024, 1,024, 512 and 1 nodes. The first four dense layers use dropout. The last layer uses linear activation to predict chronological age. A single layer (25 nodes) neural network is trained on sex and ethnicity, and its output is concatenated with the output of the second dense layer before being inputted in the third dense layer. All layers, except for the last one, use batch regularization, kernel and bias regularization, and use ReLU as their activation function.

We used Adam ^56^ as the compiler for all the time series models.

#### Images

##### Convolutional Neural Networks Architectures

We used transfer learning ^57–59^ to leverage two different convolutional neural networks ^60^ [CNN] architectures pre-trained on the ImageNet dataset ^61–63^ and made available through the python Keras library ^64^: InceptionV3 ^65^ and InceptionResNetV2 ^66^. We considered other architectures such as VGG16 ^67^, VGG19 ^67^ and EfficientNetB7 ^68^, but found that they performed poorly and inconsistently on our datasets during our preliminary analysis and we therefore did not train them in the final pipeline. For each architecture, we removed the top layers initially used to predict the 1,000 different ImageNet images categories. We refer to this truncated model as the “base CNN architecture”.

We added to the base CNN architecture what we refer to as a “side neural network”. A side neural network is a single fully connected layer of 16 nodes, taking the sex and the ethnicity variables of the participant as input. The output of this small side neural network was concatenated to the output of the base CNN architecture described above. This architecture allowed the model to consider the features extracted by the base CNN architecture in the context of the sex and ethnicity variables. For example, the presence of the same physical activity feature can be interpreted by the algorithm differently for a male and for a female. We added several sequential fully connected dense layers after the concatenation of the outputs of the CNN architecture and the side neural architecture. The number and size of these layers were set as hyperparameters. We used ReLU ^69^ as the activation function for the dense layers we added, and we regularized them with a combination of weight decay ^70,71^ and dropout ^72^, both of which were also set as hyperparameters. Finally, we added a dense layer with a single node and linear activation to predict age.

##### Compiler

The compiler uses gradient descent ^73,74^ to train the model. We treated the gradient descent optimizer, the initial learning rate and the batch size as hyperparameters. We used mean squared error [MSE] as the loss function, root mean squared error [RMSE] as the metric and we clipped the norm of the gradient so that it could not be higher than 1.0 ^75^.

We defined an epoch to be 32,768 images. If the training loss did not decrease for seven consecutive epochs, the learning rate was divided by two. This is theoretically redundant with the features of optimizers such as Adam, but we found that enforcing this manual decrease of the learning rate was sometimes beneficial. During training, after each image has been seen once by the model, the order of the images is shuffled. At the end of each epoch, if the validation performance improved, the model’s weights were saved.

We defined convergence as the absence of improvement on the validation loss for 15 consecutive epochs. This strategy is called early stopping ^76^ and is a form of regularization. We requested the GPUs on the supercomputer for ten hours. If a model did not converge within this time and improved its performance at least once during the ten hours period, another GPU was later requested to reiterate the training, starting from the model’s last best weights.

### Training, tuning and predictions

We split the entire dataset into ten data folds. We then tuned the models built on scalar data, on time series, on images and on videos using four different pipelines. For scalar data-based models, we performed a nested-cross validation. For time series-based and images-based models, we manually tuned some of the hyperparameters before performing a simple cross-validation. We describe the splitting of the data into different folds and the tuning procedures in greater detail in the Supplementary.

### Interpretability of the machine learning predictions

To interpret the models, we used the regression coefficients for the elastic nets, the feature importances for the GBMs, a permutation test for the fully connected neural networks, and attention maps (saliency and Grad-RAM) for the convolutional neural networks (Supplementary Methods).

### Ensembling to improve prediction and define aging dimensions

We built a three-level hierarchy of ensemble models to improve prediction accuracies. At the lowest level, we combined the predictions from different algorithms on the same aging subdimension. For example, we combined the predictions generated by the elastic net, the gradient boosted machine and the neural network on the scalar predictors (subdimension). At the second level, we combined the predictions from different subdimensions of a unique dimension. For example, we combined the models built on the seven subdimensions (one scalar-based model, two time series-based models and four image-based models) of the “Full week” dimension into an ensemble prediction. Finally, at the highest level, we combined the predictions from the two physical activity dimensions into a general biological age prediction. The ensemble models from the lower levels are hierarchically used as components of the ensemble models of the higher models. For example, the ensemble models built by combining the algorithms at the lowest level for each of the Full Week subdimensions are leveraged when building the general, overarching ensemble model.

We built each ensemble model separately on each of the ten data folds. For example, to build the ensemble model on the testing predictions of the data fold #1, we trained and tuned an elastic net on the validation predictions from the data fold #0 using a 10-folds inner cross-validation, as the validation predictions on fold #0 and the testing predictions on fold #1 are generated by the same model (see Methods - Training, tuning and predictions - Images - Scalar data - Nested cross-validation; Methods - Training, tuning and predictions - Images - Cross-validation). We used the same hyperparameters space and Bayesian hyperparameters optimization method as we did for the inner cross-validation we performed during the tuning of the non-ensemble models.

To summarize, the testing ensemble predictions are computed by concatenating the testing predictions generated by ten different elastic nets, each of which was trained and tuned using a 10-folds inner cross-validation on one validation data fold (10% of the full dataset) and tested on one testing fold. This is different from the inner-cross validation performed when training the non-ensemble models, which was performed on the “training+validation” data folds, so on 9 data folds (90% of the dataset).

### Evaluating the performance of models

We evaluated the performance of the models using two different metrics: R-Squared [R^2^] and root mean squared error [RMSE]. We computed a confidence interval on the performance metrics in two different ways. First, we computed the standard deviation between the different data folds. The test predictions on each of the ten data folds are generated by ten different models, so this measure of standard deviation captures both model variability and the variability in prediction accuracy between samples. Second, we computed the standard deviation by bootstrapping the computation of the performance metrics 1,000 times. This second measure of variation does not capture model variability but evaluates the variance in the prediction accuracy between samples.

### Biological age definition

We defined the biological age of participants as the prediction generated by the model, after correcting for the bias in the residuals.

We indeed observed a bias in the residuals. For each model, participants on the older end of the chronological age distribution tend to be predicted younger than they are. Symmetrically, participants on the younger end of the chronological age distribution tend to be predicted older than they are. This bias does not seem to be biologically driven. Rather it seems to be statistically driven, as the same 60-year-old individual will tend to be predicted younger in a cohort with an age range of 60-80 years, and to be predicted older in a cohort with an age range of 60-80. We ran a linear regression on the residuals as a function of age for each model and used it to correct each prediction for this statistical bias.

After defining biological age as the corrected prediction, we defined accelerated aging as the corrected residuals. For example, a 60-year-old with a predicted age of 70 years old after correction for the bias in the residuals is estimated to have a biological age of 70 years, and an accelerated aging of ten years.

It is important to understand that this step of correction of the predictions and the residuals takes place after the evaluation of the performance of the models but precedes the analysis of the biological ages properties.

### Genome-wide association of accelerated aging

The UKB contains genome-wide genetic data for 488,251 of the 502,492 participants ^77^ under the hg19/GRCh37 build.

We used the average accelerated aging value over the different samples collected over time (see Supplementary - Models ensembling - Generating average predictions for each participant). Next, we performed genome wide association studies [GWASs] to identify single-nucleotide polymorphisms [SNPs] associated with accelerated aging using BOLT-LMM ^30,31^ and estimated the the SNP-based heritability for each of our biological age phenotypes, and we computed the genetic pairwise correlations between dimensions using BOLT-REML ^32^. We used the v3 imputed genetic data to increase the power of the GWAS, and we corrected all of them for the following covariates: age, sex, ethnicity, the assessment center that the participant attended when their DNA was collected, and the 20 genetic principal components precomputed by the UKB. We used the linkage disequilibrium [LD] scores from the 1,000 Human Genomes Project ^78^. To avoid population stratification, we performed our GWAS on individuals with White ethnicity.

#### Identification of SNPs associated with accelerated aging

We identified the SNPs associated with accelerated aging using the BOLT-LMM ^30,31^ software (p-value of 5e-8). The sample size for the genotyping of the X chromosome is one thousand samples smaller than for the autosomal chromosomes. We therefore performed two GWASs for each aging dimension. (1) excluding the X chromosome, to leverage the full autosomal sample size when identifying the SNPs on the autosome. (2) including the X chromosome, to identify the SNPs on this sex chromosome. We then concatenated the results from the two GWASs to cover the entire genome, at the exception of the Y chromosome.

We plotted the results using a Manhattan plot and a volcano plot. We used the bioinfokit ^79^ python package to generate the Manhattan plots. We generated quantile-quantile plots [Q-Q plots] to estimate the p-value inflation as well.

#### Heritability and genetic correlation

We estimated the heritability of the accelerated aging dimensions using the BOLT-REML ^32^ software. We included the X chromosome in the analysis and corrected for the same covariates as we did for the GWAS.

We annotated the significant SNPs with their matching genes using the following four steps pipeline. (1) We annotated the SNPs based on the rs number using SNPnexus ^80–84^. When the SNP was between two genes, we annotated it with the nearest gene. (2) We used SNPnexus to annotate the SNPs that did not match during the first step, this time using their genomic coordinates. After these two first steps, 30 out of the 9,697 significant SNPs did not find a match. (3) We annotated these SNPs using LocusZoom ^85^. Unlike SNPnexus, LocusZoom does not provide the gene types, so we filled this information with GeneCards ^86^. After this third step, four genes were not matched. (4) We used RCSB Protein Data Bank ^87^ to annotate three of the four missing genes. One gene on the X chromosome did not find a match (position 56,640,134).

### Non-genetic correlates of accelerated aging

We identified non-genetically measured (i.e factors not measured on a GWAS array) correlates of each aging dimension, which we classified in six categories: biomarkers, clinical phenotypes, diseases, family history, environmental, and socioeconomic variables. We refer to the union of these association analyses as an X-Wide Association Study [XWAS]. (1) We define as biomarkers the scalar variables measured on the participant, which we initially leveraged to predict age (e.g. blood pressure, Table S3). (2) We define clinical phenotypes as other biological factors not directly measured on the participant, but instead collected by the questionnaire, and which we did not use to predict chronological age. For example, one of the clinical phenotypes categories is eyesight, which contains variables such as “wears glasses or contact lenses”, which is different from the direct refractive error measurements performed on the patients, which are considered “biomarkers” (Table S6). (3) Diseases include the different medical diagnoses categories listed by UKB (Table S9). (4) Family history variables include illnesses of family members (Table S11). (5) Environmental variables include alcohol, diet, electronic devices, medication, sun exposure, early life factors, medication, sun exposure, sleep, smoking, and physical activity variables collected from the questionnaire (Table S12). (6) Socioeconomic variables include education, employment, household, social support and other sociodemographics (Table S15). We provide information about the preprocessing of the XWAS in the Supplementary Methods.

## Data Availability

https://www.multidimensionality-of-aging.net/

https://www.dropbox.com/s/59e9ojl3wu8qie9/Multidimensionality_of_aging-GWAS_results.zip?dl=0

## Acknowledgments

We would like to thank Raffaele Potami from Harvard Medical School research computing group for helping us utilize O2’s computing resources. We thank HMS RC for computing support. We also want to acknowledge UK Biobank for providing us with access to the data they collected. The UK Biobank project number is 52887.

## Author Contributions

**Alan Le Goallec:** (1) Designed the project. (2) Supervised the project. (3) Predicted chronological age from images. (4) Computed the attention maps for the images. (5) Generated preliminary results for the chronological age predictors built on scalar features. (6) Ensembled the models, evaluated their performance and computed biological age. (7) Performed the genome wide association studies. (8) Designed the website. (9) Wrote the manuscript.

**Sasha Collin:** (1) Preprocessed the data: generated the scalar features, preprocessed the time series and generated the images. (2) Predicted chronological age from the time series datasets. (3) Computed the attention maps for the time series.

**M’Hamed Jabri:** (1) Helped preprocess the data, engineer features and generate recurrence plots. (2) Generated preliminary results for the prediction of chronological age from scalar features.

**Samuel Diai:** (1) Predicted chronological age from scalar features. (2) Wrote the python class to build an ensemble model using a cross-validated elastic net. (3) Performed the X-wide association study. (4) Implemented a first version of the website https://www.multidimensionality-of-aging.net/.

**Théo Vincent:** (1) Website data engineer. (2) Implemented a second version of the website https://www.multidimensionality-of-aging.net/.

**Chirag J. Patel:** (1) Supervised the project. (2) Edited the manuscript. (3) Provided funding.

