## Supplementary Information for "Predicting age from 100,000 one week-long 100Hz wrist accelerometer records of physical activity"

#### Table of contents

**Table of contents** 1

**Discussion** 2

Comparison between our models and the literature 2

Physical activity: daily life 2

Physical activity: Gait analysis 5

**Methods** 6

Hardware 6

Software 6

Training, tuning and predictions 7

Data splitting 7

Scalar data 7

Nested cross-validation 7

Bayesian hyperparameters optimization 8

Example 8

Images 11

Hyperparameters tuning upstream of the cross-validation 11

Cross-validation 14

Cross-validation example 14

Time series 15

Hyperparameters tuning upstream of the cross-validation 16

Tuning of the seed using a cross-validation 17

Generating average predictions for each participant 17

Interpretability of the predictions 19

Scalar data-based predictors 19

Time series, image- and video-based predictors 19

Non-genetic correlates of accelerated aging 23

Imputation of the non-genetic X-variables 23

X-Wide Association Studies 25

**Supplementary Figures** 26

**Supplementary Tables** 38

### Discussion

#### Comparison between our models and the literature

A summary of our comparison between our models and the literature can be found in Table S19.

##### Physical activity: daily life

Other investigators have built a survival model on UKB's wrist accelerometer data [1] predicting all-cause mortality, but we are, to our knowledge, the first to predict chronological age using this dimension/data modality. We compare our performance to two chronological age predicting models built on NHANES's wrist accelerometer data. These two publications are the ones from which we adapted the CNN architectures to build our own pipeline (see Methods - Machine learning algorithms - Time series - Physical activity) [2,3].

First, Pyrkov et al. predicted chronological age on 7,454 samples with an age range of 6-84 years and obtained a $R^2$ value of approximately 56% (Pearson correlation: 75%) and a RMSE of 14.0 years using a CNN [2].

Second, Rahman and Adjeroh predicted chronological age on 4,268 samples with an age range of 18-84 years with a RMSE of 15.74 years [3]. Rahman and Adjeroh also reported $R^2$ values as high as 0.93. We want to be very careful in raising this point, but we suggest there might have been an error in the code when computing the $R^2$ values for some of the models, and we provide four pieces of evidence to support this suggestion. (1) The order of magnitude for the $R^2$ values does not match the other metrics (Table 10 in Rahman et al's publication). For example, the training RMSE, correlation and $R^2$ values for the 1D CNN model are respectively 18.81 years, 52% and 88%. For a linear regression, the training $R^2$ value is the square of the training Pearson correlation, which gives us an order of magnitude of 27% rather than the 88% reported. (2) The testing MAE (15.49 years), RMSE (18.81 years) and correlation (45%) values reported show that the prediction accuracy on the testing set is lower than on the training set, as expected. However, the testing $R^2$ value reported is 93%, higher than the training $R^2$ value (88%). Similar anomalies can be observed for the other architectures (DNN, ConvLSTM, CNN+LSTM). (3) Sagers et al. reported a similar testing $R^2$ value of 92% on NHANES using laboratory biomarkers[4]. They, however, reported a corresponding MAE value of 4.76 years for a comparable age range that was more than three times lower than the testing MAE reported by Rahman and Adjeroh for their 1D CNN (MAE: 15.49 years, $R^2$=93%). (4) Rahman and Adjeroh themselves noticed the discrepancy in their results: “While 1D CNN has the best $R^2$ (0.93) followed by the DNN network ($R^2$=0.89), for MAE and correlation ConvLSTM* and CNN+LSTM model performed better.”. Based on the four aforementioned arguments, we will approximate Rahman et al's testing $R^2$ value by the square of their best model's testing correlation ($0.62^2$=38.4%) to compare the performance of their models with ours.

Our ensemble model predicted chronological age using wrist accelerometer data with a RMSE of 4.71±0.1 years and a $R^2$ value of 63.5±0.2%. Our RMSE is therefore approximately three times smaller than the RMSEs reported by Pyrkov et al. (RMSE=14.0) and Rahman and

Adjeroh (RMSE=15.74), but our better performance is in part driven by the narrower age range covered by UKB (UKB: 43-79 years; Pyrkov et al.: 6-84 years; Rahman and Adjeroh: 18-84 years). Our model also outperformed the two NHANES based models in terms of $R^2$ values (our model: 63.5±0.2%; Pyrkov et al.: 56%; Rahman and Adjeroh: 38.4%) despite being built on a narrower age range.

We hypothesize that four factors can explain our better performance. (1) The sample size we leveraged from the UKB dataset after preprocessing and filtering the samples is twelve to twenty times larger the sample sizes that Pyrkov et al. and Rahman and Adjeroh could extract from the NHANES dataset (UKB: 90,000-100,000, NHANES: 4,268-7,454). (2) Unlike UKB, NHANES accelerometer data is a frequency measure that does not capture the absolute value of the acceleration. UKB data is also available at two different sampling frequencies (0.2 Hz and 100 Hz), which we both leveraged. (3) We found that our best predictive non-ensemble model was the GBM we trained on 2,271 scalar features ($R^2$=59.4±0.2%, RMSE=5.11±0.1 years). In contrast, our best non-ensemble model built on time series gave a $R^2$ value of 41.8±0.3% and a RMSE of 6.11±0.1 years. The models built by Pyrkov et al. and Rahman and Adjeroh rely on time series data compared to our scalar based model. (4) Building ensemble models increased our $R^2$ value from $R^2$=59.4±0.2% to 63.5±0.2%. Both Pyrkov et al. and Rahman and Adjeroh built several models but since, to our knowledge, they did not create an ensemble, we report the performances of their non-ensembled models. (5) We built both the CNN architecture designed by Pyrkov et al. and the ConvLSTM* architecture designed by Rahman and Adjeroh, which is their best performing architecture. We found that the architecture designed by Pyrkov et al. outperformed the architecture designed by Rahman and Adjeroh on the UKB dataset, which explains in part why we report higher prediction performances than Rahman and Adjeroh. Our better RMSE values than Pyrkov et al. (6.11±0.1 years vs. 14.0 years) despite using the same CNN architecture is likely driven by the narrower UKB's age, as well as reasons (1) and (2) mentioned above (sample size and difference in time series types).

#### Physical activity: Gait analysis

Gait changes as one gets older [5] and several research teams predicted chronological age from the gait [6–14]. Lu and Tan analyzed the gait of 1,870 silhouette gaits collected from participants aged 20-30 years to predict chronological age with a MAE of 3.02 years [9]. Makihara et al. analyzed 1,728 gait videos collected from participants aged 2-94 years and predicted chronological age with a MAE of 8.2 years [11]. Riaz et al. attached integrated measurement units to the lower back of 26 participants (mean age: 48.1 years old; standard deviation: 12.7 years) and recorded their gait. They then trained a random forest on the features extracted from these records and classified the participants into three age categories (age < 40 years, 40 years < age < 50 years and age > 50 years) with 88.82% accuracy [12]. Hoffmann et al. recorded the gait of 142 participants aged 13-82 years as they walked over a sensor floor. They then built a neural network and predicted chronological age with a MAE of 9.55 years and a $R^2$ value of 37.21% [14]. Li et al. built a support vector machine on features extracted from 63,846 gait energy image[15] recordings of participants aged 2-90 years. They predicted chronological age with a MAE of 6.78 years [13].

We analyzed the gait of the 77,269 UKB participants using wrist accelerometer recordings and predicted chronological age with a $R^2$ of 32.3±0.2% and a RMSE of 5.69±0.01 years. Only Hoffmann et al. reported a $R^2$ value (37.21%)[14], larger than the one we obtained (32.3±0.2%), which can be explained by the wider age range covered by their dataset. The RMSE we obtained is lower than the MAE values reported by the aforementioned publications, which can be explained by the narrower age range covered by UKB (37-82 years). Comparing the performance of our model with the ones reported in the literature is therefore difficult, but we hypothesize that the gait is more efficiently analyzed using gait videos than wrist accelerometer recordings and that Li et al.'s predictor is therefore likely more accurate than the one we built [13].

### Methods

#### Hardware

We performed the computation for this project on Harvard Medical School's compute cluster, with access to both central processing units [CPUs] and general processing units [GPUs] (Tesla-M40, Tesla-K80, Tesla-V100) via a Simple Linux Utility for Resource Management [SLURM] scheduler.

#### Software

We coded the project in Python [16] and used the following libraries: NumPy [17,18], Pandas [19], Matplotlib [20], Plotly [21], Python Imaging Library [22], SciPy [23–25], Scikit-learn [26], LightGBM [27], XGBoost [28], Hyperopt [29], TensorFlow 2 [30], Keras [31], Keras-vis [32], iNNvestigate [33]. We used Dash [34] to code the website on which we shared the results. We set the seed for the os library, the numpy library, the random library and the tensorflow library to zero.

### Training, tuning and predictions

#### Data splitting

We split the 676,787 samples into ten data folds, while keeping all samples from the same participant in the same fold. To ensure this, we split the 502,211 participants' ids (referred to by UKB as "eid") into ten different buckets of the same size.

#### Scalar data

##### Nested cross-validation

Cross-validation is a method to tune the regularization of models and prevent overfitting [35]. For the models inputting scalar data (Figure 1A in green), we tuned the hyperparameters and generated a testing prediction for each sample using a nested 10x9-folds cross-validation. We refer to the two nested cross-validations as the "outer" and the "inner" cross-validations. The outer-cross validation is used to generate an unbiased testing prediction for each sample, as opposed to a simple split of the data into a "training+validation" set on one hand, and a testing set on the other hand, which would only generate a testing prediction for one tenth of the dataset. The inner cross-validation is used to tune the hyperparameters more precisely, leveraging the full inner cross-validation dataset as a validation set, as opposed to a simple data split of the "training+validation" dataset into a training and a validation sets, which would only use one data fold as the validation set to estimate the performance associated with a specific combination of hyperparameters. The nested cross-validation is illustrated in Table S21.

#### Bayesian hyperparameters optimization

To tune the hyperparameters, we used the Tree-structured Parzen Estimator Approach [36] [TPE] of the hyperopt python package [37]. TPE is a sequential Bayesian hyperparameters optimization method that iteratively suggests the next most promising hyperparameters combination as a function of the hyperparameters combinations that have already been tested, by building a probabilistic representation of the objective function. We set the number of iterations to 30. For each model, 30 different hyperparameter combinations are iteratively tested before selecting the best performing one. The hyperparameters names and their ranges defining the hyperparameters space can be found in Table S39. It might be of interest to other researchers that we initially tuned the hyperparameters using a random search [38] with the same number of iterations, and we did not observe a significant improvement in the model's performance after implementing the Bayesian hyperparameters optimization.

#### Example

For the sake of clarity, let us walk through a concrete example, which is illustrated in Table S41. Suppose we want to generate unbiased predictions for every sample in a dataset using an elastic net. First, let us generate the testing prediction for the data fold F9, which is performed by the first fold of the outer cross-validation (outer cross-validation fold 0). We select the data fold F9 out of the ten data folds as the testing fold, and we select the remaining nine data folds as "training+validation" folds for the inner cross-validation. We scale and center the target (age) and the predictors using the mean and standard deviation values of the variables on the "training+validation" dataset. We then enter the first inner-cross validation.

For the first inner cross-validation fold, we select the data fold F8 as the validation set, and the remaining eight “training+validation” data folds as the training set. We re-scale and center age and the predictors in the training and the validation sets using the mean and standard deviation values of the training set. We train the model on the eight training data folds with the first hyperparameters combination sampled by the TPE algorithm (one value for alpha and one value for l1_ratio) and generate validation predictions on the validation fold (data fold F8), which we unscale. This completes the first of the nine inner cross-validation folds (Inner CV fold 0). We then permute the nine inner data folds. We scale the age and the predictors using the mean and standard deviation computed on the new training set. Then we train the model with the same first combination of hyperparameters on eight data folds, leaving aside the data fold F9 (still being used as the testing set for the outer cross-validation) and the data fold F7 (now being used as the validation set for the inner cross-validation). We then use the new trained model to generate validation predictions on the data fold F7, which we unscale. This completes the second of the nine inner-cross validation folds (Inner CV fold 1). We then reiterate these inner permutation and training processes seven more times, until every data fold in the nine “training+validation” data folds is used as the validation set once. At this point, we concatenate the validation predictions from these nine validation folds to obtain the overall validation predictions associated with the first hyperparameters combination, and compute the associated performance metric (e.g. RMSE). This completes the inner-cross validation for the first hyperparameters combination.

We then perform the same 9-folds inner cross-validation, this time with the second hyperparameters combination suggested by the TPE algorithm. We iterate this process 28 more times, until 30 different hyperparameters combinations have iteratively been tested. Next, we select the hyperparameter combination that yielded the best validation performance (e.g. minimum RMSE), and we retrain a model on the whole nine “training+validation” data folds (all data folds except for data fold #1), using this best performing hyperparameters combination. This completes the first inner cross-validation.

We then use the model to generate unbiased predictions on the unseen testing set (data fold F9) and record these predictions. By anticipation for the ensembling algorithm (see Methods - Models ensembling) we also need to compute validation predictions on the data fold F8. We do this by training a model on all the data folds aside from the validation fold (data fold F8) and the testing fold (data fold F9), with the selected hyperparameters combination. We then use this trained model to compute predictions on the validation fold (data fold F8) and record these predictions, after unscaling them. This completes the first of the ten outer cross-validation folds (outer cross-validation 0).

We then complete the second outer cross-validation fold (outer cross-validation 1), this time using the data fold F8 as the testing dataset, to obtain unbiased testing predictions on this data fold, as well as validation predictions on the data fold F7. We reiterate the process eight more times to obtain the testing and validation predictions on the remaining data folds. We then concatenate the testing predictions from the ten data folds to obtain our final testing predictions for the model. Similarly, we concatenate the validation predictions from the ten data folds to obtain our final testing predictions for the model, which will later be used during ensemble models building and model selection (see Methods - Models ensembling).

The final validation and testing predictions for each data fold are therefore not necessarily associated with the same hyperparameters combination. It is also important to notice that we performed a single outer cross-validation, but that we performed a separate inner-cross validation for each outer cross-validation fold (hence the word “nested”), for a total of ten inner cross-validations per outer cross-validation fold.

#### Images

##### Hyperparameters tuning upstream of the cross-validation

The hyperparameters we tuned were the number of added fully connected dense layers, the number of nodes in these layers, their activation function, the optimizer, the initial learning rate, the weight decay, the dropout rate, the data augmentation amplitude and the batch size. Repeatedly tuning the values of the hyperparameters for different deep neural networks architectures and on the different cross-validation folds would have been prohibitively time and resource consuming. Instead, we sequentially explored how each hyperparameter was affecting the training and validation performances for a single architecture (InceptionV3) on a single cross validation fold (fold #0, see Methods - Training, tuning and predictions - Images - Cross-validation for the detailed description of the cross-validation). We then extrapolated the hyperparameter values to the other architectures, datasets and cross-validation folds. The hyperparameters combinations tested during the tuning can be found in Table S22.

First, we maximized the batch size for each architecture. The maximum number of images per batch depends on the memory of the GPU and the size of the architecture, which itself depends on the dimensions of the image. We used a batch size of 32 for InceptionV3 and 8 for InceptionResNetV2.

Then, we tested the learning rates, including 1e-6, 1e-5, 1e-4, 1e-3, 1e-2 and 1e-1. We observed that learning rates larger than 1e-4 prevented the model from converging for some runs. Second, we did not observe significant differences between the results obtained with learning rates smaller than 1e-4. We therefore set the initial learning rate to be 1e-4 for all models to shorten the time to convergence while ensuring that the learning rate was small enough to allow convergence and the finding of a local minima for the loss function.

Then we tested three different optimizers to perform the gradient descent: Adam [39], Adadelta [40] and RMSprop [41]. We did not observe any significant differences between the optimizers, so we set the optimizer to be Adam.

We then added different numbers of fully connected layers between the base CNN and side CNN's concatenated outputs and the final activation layer. We set the number of nodes to be 1,024 in the first added layer and then decreased the number of nodes by a factor of two for each successive layer. For example, if we added three fully connected layers, the number of nodes was 1024, 512 and 256. We added zero, one and five layers. We did not observe significant differences in the performance of the different architectures, so we set the number of fully connected layers to one.

We then tested powers of two from 16 to 2,048 as the number of nodes in this single layer. We did not observe significant differences between these architectures, so we set the number of nodes to be 1,024 to keep the number close to the initial number of nodes in the imported CNN architectures, as these were initially used to perform classification between 1,000 categories.

We tested two different activation functions for the activation functions of the fully connected layers we added in the side neural network and before the final linear layer. We did not observe any significant differences between the rectified linear units [ReLU] [42] and the scaled exponential linear units [SELU] [43] as activation functions, so we used the more common ReLU.

We then tested different levels of data augmentation. We introduced a hyperparameter that we called “data augmentation factor”. The data augmentation factor modulates the amount of variation introduced by the data augmentation, while preserving the ratio between the different transformations. For example, a data augmentation factor of one is equivalent to the default data augmentation (see Preprocessing - Data augmentation - Images), but a data augmentation factor of two will double the ranges of the possible values sampled and the expected values for the vertical shift, the horizontal shift, the rotation and the zoom on the original images. We tested the following values for the data augmentation factor: 0, 0.1, 0.5, 1, 1.5 and 2. We found that different values for the data augmentation factor hyperparameter yielded similar results, as long as the data augmentation factor was not zero. We therefore set the data augmentation factor to be one when training the final models.

We then tuned the dropout rate for the fully connected layers we added. We tested the following values: 0, 0.1, 0.25, 0.3, 0.5, 0.75, 0.9 and 0.95. We observed that a dropout rate of 0.95 led to underfitting and that smaller values reduced overfitting on the training set but without improving the validation performance. As a consequence, we used a dropout rate of 0.5.

Finally, we tuned the weight decay. We tested the following values: 0, 0.1, 0.2, 0.3, 0.4, 0.5, 1, 5, 10 and 100. For the larger datasets, we found that weight decay values as low as 0.4 could lead to underfitting. We found that lower weight decay values reduced overfitting on the training set without significantly improving the validation performance. We set the weight decay to 0.1. Altogether, we found that hyperparameter tuning had little effect on the validation performance as long as extreme hyperparameters values were not selected.

##### Cross-validation

Training deep convolutional neural networks on images and videos is too time and resource consuming to perform a nested cross-validation. Therefore, we tuned the hyperparameters during the preliminary analysis, as described above. After hyperparameters tuning, we performed a simple outer cross-validation to obtain a testing prediction for each sample of the datasets, but we replaced the inner cross-validation with a simple split between the training fold and the validation fold (Table S23). Although the hyperparameters were already tuned, a validation set was still required for two reasons: (1) to perform early stopping [44], a form of regularization. (2) to generate a set of validation predictions that are necessary for efficient ensemble building (see Methods - Models ensembling) and model selection. During the cross-validation, we scaled and centered the target variable (chronological age) as well as the side predictors (sex and ethnicity) around zero with a standard deviation of one, using the training summary statistics. Scaling the target and the input helps prevent the issues of exploding and vanishing gradients [45,46].

##### Cross-validation example

For the sake of clarity, let us walk through an example. Let us say that we want to generate unbiased predictions for every sample in a dataset using a CNN. First, we select the data fold #0 as the validation set, the data fold #1 as the testing set, and the remaining data folds (#2-9) as the training set. Then we scale and center the target (age), and the side predictors (sex and ethnicity) using the training mean and standard deviation: for each of the variables, we subtract the training mean to the variable on both the training, the validation and the testing set, and we divide it by the training standard deviation. We then train the model on the training set until convergence and select the architecture's parameters (also known as "weights") associated with the epoch that yielded the lowest validation RMSE. We then use the optimal weights to generate validation predictions for the data fold #0 and testing predictions on the data fold #1. Finally, we unscale the validation and testing predictions by multiplying them by the initial age training standard deviation before adding the initial age training mean to them. This completes the first cross-validation fold.

We then reiterate the process, this time using the data fold #1 as the validation set, the data fold #2 as the testing set, and the remaining data folds (#0 and #3-9) as the training set. We use the optimized weights to generate the validation predictions on the data fold #2, and the testing predictions on the data fold #3. We unscale the validation and testing predictions. This completes the second cross-validation fold. We reiterate the process eight more times to complete the cross-validation. We then concatenate the validation predictions from the ten data folds to obtain the final validation predictions, and the testing predictions from the ten data folds to obtain the final testing predictions.

#### Time series

We tuned the time series models in two steps. (1) We tuned the hyperparameters using a single cross-validation fold. (2) We tuned the seed and performed early stopping (patience=40) using a simple cross-validation, to generate a testing sample size for every sample. These two steps are described in detail below.

#### Hyperparameters tuning upstream of the cross-validation

The hyperparameters we tuned were the number of convolutional layers in the architecture, the number of nodes in the dropout rate and the strength of the kernel and the bias regularizations. The tuned the hyperparameters using a single cross-validation fold. Specifically, we used the data fold #0 as validation set and the data folds #2-9 as training set. We used the same pipeline as for the images and the videos, with two differences. (1) Because the training of models built on time series was significantly faster than the training of models built on images or videos, we used a grid search to tune the hyperparameters rather than tune them sequentially. (2) We scaled the input differently. For the models built on the raw acceleration data across the full week, we did not scale the data because we adapted architectures from publications in which the input has not been scaled. For the models built on features extracted from acceleration data and three-dimensional walking data, we normalized every sample separately by dividing each lead by the absolute value of its maximal value for the sample. We then used this maximum value as a scalar predictor which we refer to in this paper as “scaling factor”.

We tested architectures with one to ten convolutional layers for the convolutional block. The default number of filters for each convolutional layer doubled with every layer, starting from 16 and capped at 1,024. For example, the default number of filters for a nine layers deep convolutional block would be 16, 32, 64, 128, 256, 512, 1,024, 1,024 and 1,024. To allow the architecture to increase its breadth without increasing its depth, we introduced another hyperparameter which we called the “filters factor”, with a value of either one, two or four. The number of filters in each convolutional layer was the default number of filters multiplied by the filters factor value. For example, the number of filters for a three-layer deep convolutional block with a filter factor of four would be 64, 128 and 256, instead of 16, 32 and 64. For the dropout rate, we defined the hyperparameter space as eleven values uniformly spread between 0 and 50%, included. For the kernel and the bias regularizers, we defined the hyperparameter space as the absence of regularization (0) and every negative power of 10 between three (10e-3) and six (10e-6), included. We used the hyperparameters values selected on this first cross-validation fold for the nine remaining cross-validation folds. The hyperparameters values selected can be found in Table S46.

##### Tuning of the seed using a cross-validation

We observed that the hyperparameters values selected on the first cross-validation fold did not always perform as well for the other cross-validation folds. Similarly, we observed that similar hyperparameter values combinations could lead to significantly different performances on the first cross-validation fold. We hypothesized that these differences could be partly driven by different random initializations of the weights of the neural network, which led to convergence to different local minima. To mitigate this effect, we tuned the Keras seed independently for each of the ten cross-validation folds. We tested every integer between zero and nine, included, and selected the seed that yielded the best model in terms of performance on the validation set.

The cross-validation also served to generate a testing prediction on every sample and to perform early stopping, as described under the pipeline for images.

#### Generating average predictions for each participant

We generated an average prediction for each individual, reported to UKB's instance 0. We walk through an example. Let us assume a participant had two wrist accelerometer records collected from them in instances 2 and 3, respectively at age 70 and 80. Let us assume that the age predictions were respectively 64 and 78, so the residuals are respectively -6 years and -2 years, for an average of -4 years. However, we still need to take into account the bias in the residuals, defined as the difference between the participant's chronological age and the prediction. As explained in more details under Methods - Biological age definition, we observed a bias in the residuals as a function of chronological age. Participants on the younger end of the chronological age distribution tend to be predicted older than they actually are, whereas participants on the older end of the distribution tend to be predicted younger than they actually are. We need to properly account for this bias when translating a prediction from a more recent instance to an older instance. Let us assume that the average bias in the residuals for participants who are 70 and 80 years old is respectively -2 years and -4 years. After correcting for this bias, the predictions are now respectively 64-(-2)=66 and 78-(-4) = 82. Therefore, the corrected residuals for this participant are respectively -4 years and +2 years, for an average of -1 years. Finally, let us assume that the participant was 60 years old in instance 0. We will assign a single prediction of 60-1=59 years to the participant, but we still need to un-correct for the bias in residuals. Let us assume that the average bias for the residuals at age 60 is +5 years. We will assign a final prediction for the participant of 59+5=64 years.

A key point we would like to highlight here is that we did not actually correct for the bias in the residuals at this step of the pipeline. Instead, we corrected then un-corrected the predictions that we translated from different instances to the instance 0. The actual correction for the residual biases takes place when defining the biological age phenotypes (see Methods - Biological age definition).

To distinguish between raw predictions on the instance 0, and the average predictions reported to the instance 0, we created a new instance which we named instance “*”. We refer to these predictions as “participants predictions”, as opposed to “samples predictions”.

### Interpretability of the predictions

#### Scalar data-based predictors

For elastic nets, we interpreted the models using the values of the regression coefficients. Large absolute values for these coefficients means they played an important role when generating the predictions. For gradient boosted machines we used the feature importances, which are based on the number of times a tree selected each of the variables. Variables with high feature importances were selected more often and are therefore likely to play a key role in predicting chronological age. For neural networks, we estimated the importance of each feature by permuting it randomly between samples before computing the performance of the model. The score of each feature is the difference between the R-Squared value before and after the random permutations. Features whose random permutation leads to a large decrease in the model’s performance are estimated to be important predictors of chronological age.

We estimated the concordance between the three different algorithms by computing the Pearson and the Spearman correlations between their feature importances.

#### Time series, image- and video-based predictors

To interpret the CNNs built on time series, images and videos, we used saliency maps [47]. For time series and images, we coded the saliency maps using the keract python library. For videos, we generated a saliency map for each time frame using the iNNvestigate python library [33]. For each input sample, a saliency map uses the gradient of the final prediction with respect to each individual input pixel to estimate whether changing the value of this pixel would affect the prediction. Pixels for which the gradient is close to zero are not important, whereas pixels with a large gradient are estimated to be important. For videos, we computed both a saliency map for each time frame, which we stored as a .gif file, and an average saliency map over all time frames.

For images, we built a second attention map using a custom version of the Gradient-weighted Class Activation Mapping [Grad-CAM] algorithm [48] adapted to regression rather than multi-class classification: Gradient-weighted Regression Activation Mapping [Grad-RAM]. The intuition behind Grad-CAM maps is that they are similar to saliency maps [48], but instead of computing the gradient with respect to the input image, they compute it with respect to the activation of the last convolutional layer. As convolutional layers maintain the spatial organization of the input image, Grad-CAM can still identify which region of the image is driving the predictions. Because Grad-CAM does not have to backpropagate the gradient all the way back to the input image, it is considered a less noisy alternative to the saliency maps. In the same way that saliency maps need to combine the attention maps generated in the different input channels (e.g. RGB) into a single activation map, Grad-CAM must combine the attention maps generated on the different filters of the last convolutional layer. For example, the last convolutional layer for InceptionResNetV2 has 1,792 filters. Grad-CAM combines these 1,792 attention maps into a single attention map using a linear combination. In the initial Class Activation Mapping [CAM] algorithm [49], generating CAM activation maps required to retrain the model after modifying the architecture and replacing all the fully connected layers after the final convolutional layer with a global max pooling operation, which converted each filter into a scalar feature. The intuition behind this substitution was that each filter could be interpreted as detecting a specific feature, and global max pooling yielded a scalar that could be interpreted as the presence (high value) or absence (low value) of the feature anywhere on the image. The scalar values were then linearly combined and activated using the softmax function to yield the probabilities of belonging to different classes. To obtain the activation map for a specific class, the filters of the last convolution layer were linearly combined using the weights connecting the scalar features obtained after the max pooling operation to the final prediction score for that class. CAM was later improved to become Grad-CAM [48]. Grad-CAM saves the need for modifying the architecture of the model and retraining it by approximating the linear regression weight for each final convolutional filter by the mean activation gradient over the pixels of the filter. The intuition behind this approximation is that a filter's pixel is important if changing its value affects the final prediction, so a high average gradient over the pixels of the filter justifies that this filter should be given a higher weight when merging all the filters into a single attention map. To adapt Grad-CAM to our regression task we (1) computed the derivatives of the chronological age prediction rather than a class' prediction and (2) removed the ReLU activation applied to the weighted sum of the last convolutional filters, which we replaced by an absolute value. The rationale is that for (Grad-)CAM maps, we only want to highlight the regions of the picture which are associated with a high probability for the class. In contrast, for (Grad-)RAM we care as much about the regions of the input image that can strongly increase the chronological age prediction as about the regions that can strongly decrease it. Because the filters in the last convolutional layer are the result of the processing of the input image by several convolutional layers with possibly negative weights, the sign of the last convolutional layer's pixels and regression weights cannot be linked to either accelerated aging or decelerated aging, only to the magnitude of the shift that would affect the prediction if each region of the input image was modified. Regression Activation Mapping (RAM) was mentioned as a possible extension of CAM in the original CAM publication [49] and has been used to interpret models CNNs built on retinal images [50] and cortical surfaces [51], but we are to our knowledge the first to describe the generalization of Grad-CAM to a regression task. One notable difference between our implementation and Wang and Yang.'s implementation [50] is that we are taking the absolute value of the final attention map, as mentioned above. We found that not taking the absolute value led to misleading attention maps for participants with high chronological age predictions. The attention map highlights important areas with negative values, which are therefore depicted in blue, a color otherwise associated with unimportant regions in traditional CAMs. Inversely, regions on the input image for which the attention map has a slight positive value are spuriously considered to be the most important and are highlighted in red. We therefore advise that RAM or Grad-RAM be implemented using an absolute value. We coded Grad-RAM using the get_activations and get_gradients_of_activations functions of the keract python library.

It is important to understand that unlike the feature importances described under "Scalar data-based predictors", which describe the model itself, attention maps are sample specific. In other words, they can be used to explain which features drove the predictions for a specific inputted sample but cannot provide an explanation for the way the model is performing predictions in general.

For each aging subdimension, we generated the attention maps for the best performing CNN architecture. We selected representative samples for which we computed the different attention maps. We computed attention maps for the two sexes (female and male), for three age ranges (ten youngest ages, ten middle ages and ten oldest ages of the chronological age distribution) and for three aging rates (accelerated agers, normal agers, decelerated agers). For each intersection of the three categories listed above, we selected the ten most representative samples (e.g. the ten most accelerated agers among young males). The figures in this paper only present the first, most representative of these ten samples. The complete set of samples can be found on the website.

#### Non-genetic correlates of accelerated aging

Unlike DNA, biomarkers, phenotypes, diseases, family history, environmental variables and socioeconomics can change over life. As a consequence, we compared each biomarker, phenotype and environmental variable with the accelerated aging of the participant at the time the exposure was measured and we used the “Samples predictions”, as opposed to the “Participants predictions” that we used for the identification of genetic correlates (see Methods - Models ensembling - Generating average predictions for each participant).

##### Imputation of the non-genetic X-variables

Most X-variables were not collected on all four instances. Additionally, no X-variables were collected at the same time as the accelerometer data was collected. To identify the non-genetic correlates of accelerated aging, we had to impute the values of the X-variables for the ages of the participants for which they were not available. We considered two imputation methods, which we refer to as the “cross-sectional” and the “longitudinal” imputations.

For the cross-sectional imputation, we computed a linear regression for each X variable as a function of age, adjusting for sex. We then used the slope of the linear regression to extrapolate the value of the XWAS variable at different ages.

For the longitudinal imputation, we first selected, for each X variable, all the participants that had at least two measures taken for this X variable. We then performed a linear regression for each participant. We then averaged the slope of the linear regressions over all the participants of the same sex. Finally, we used this slope to extrapolate the value of the XWAS variable at different ages for all participants depending on their sex, in the same way we did it for the cross-sectional imputation.

It is important to notice that for both the cross-sectional imputation and the longitudinal imputation, data can only be imputed when the XWAS variable has been measured at least once for the participant. This raw measure is then used to extrapolate which value the X variable was likely taking a couple years earlier and/or later.

The advantage of the cross-sectional imputation is larger sample sizes. The advantage of the longitudinal method is that it corrects for generational effects. For example, old people have shorter legs than young people on average [52]. This is not because human legs shrink as we grow older. Instead, people who are old today already had shorter legs when they were young. If the cross-sectional regression is used to impute the length of the participants on instances where it was not measured, it will spuriously assign smaller values to the older samples. In contrast, the longitudinal regression learns the regression coefficient by comparing each participant to themselves as they age and will therefore not capture the generational effect.

When used to predict the participants legs' length, it will impute constant values over time. To evaluate which of the two imputation methods should be preferred, we used them to predict X-variables for which we knew the actual values and computed the R-Squared values associated with the predictions. We found that, even with sample sizes as small as 200 samples, longitudinal imputation outperformed cross-sectional imputation. We therefore used longitudinal imputation.

#### X-Wide Association Studies

First, we tested for associations in an univariate context by computing the partial correlation between each X-variable and accelerated aging. To compute the partial correlation between an X-variable and an aging, we followed a three steps process. (1) We ran a linear regression on each of the two variables, using age, sex and ethnicity as predictors. (2) We computed the residuals for the two variables. (3) We computed the correlation between the two residuals and the associated p-value if their intersection had a sample size of at least ten samples. We used a threshold for significance of 0.05 and corrected the p-values for multiple testing using the Bonferroni correction. We plotted the results using a volcano plot. We refer to this pipeline as an X-Wide Association study [XWAS].

In the supplementary tables and the results, we rank the X-variables subcategories by decreasing percentage of variables associated with accelerated aging (note that the ranking is therefore biased towards categories with fewer variables). For each subcategory, we list the three most associated variables, based on the absolute value of the correlation coefficient. For the exhaustive list, please refer to https://www.multidimensionality-of-aging.net/xwas/univariate_associations.

### Supplementary Figures

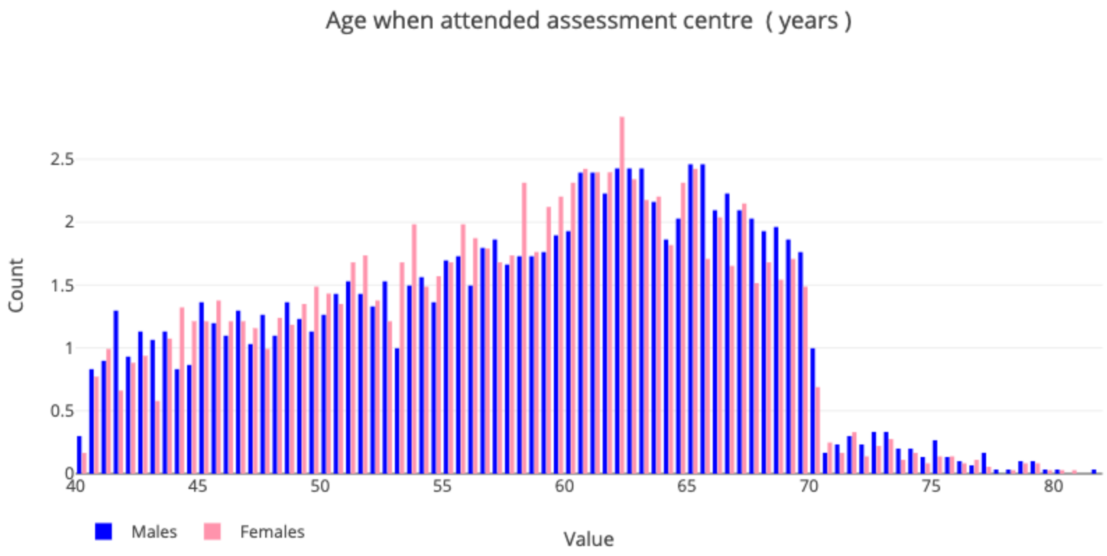

**Figure S1: Demographics of the UK Biobank cohort**

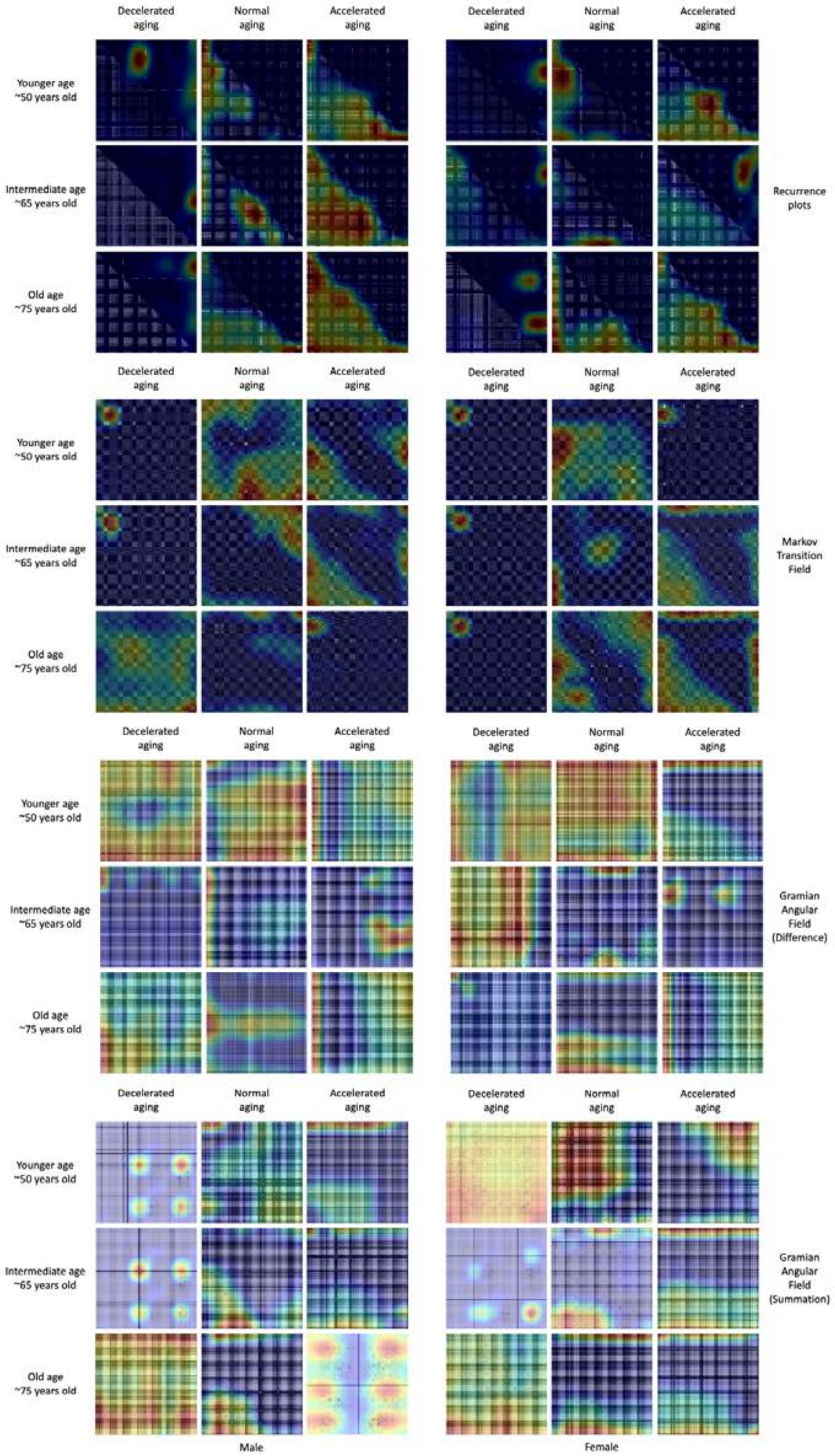

**Figure S2: Attention map samples for the images derived from the wrist accelerometer records**

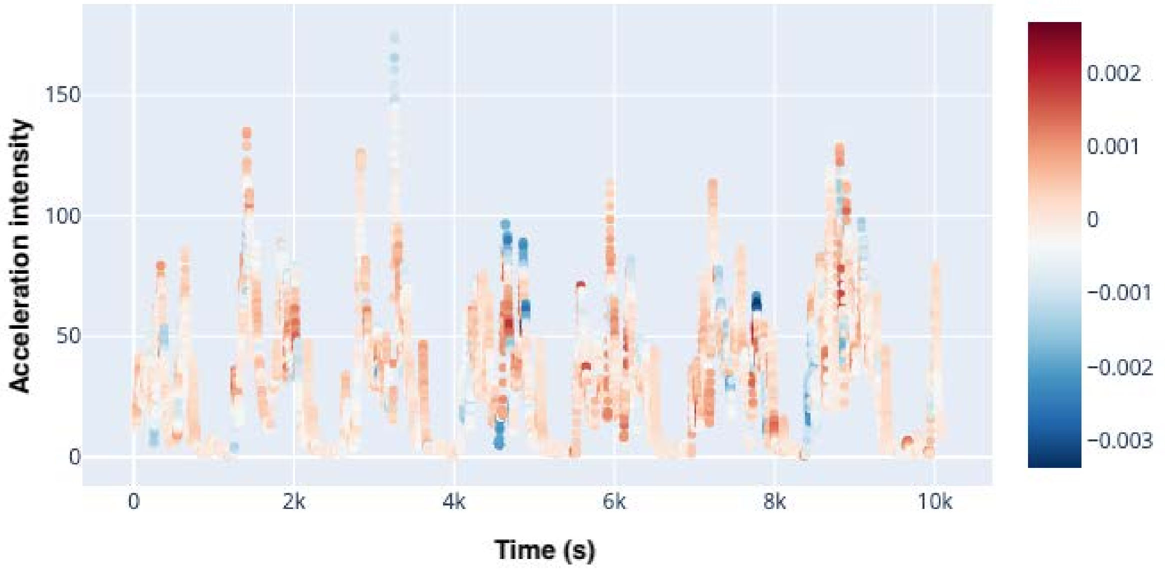

**Figure S3: Attention map example for physical activity time series - Full week**

The participant is a correctly predicted 60-65-year-old male. Red data points represent time steps for which a higher value would increase the chronological age prediction, and blue data points represent time steps for which a higher value would decrease the chronological age prediction.

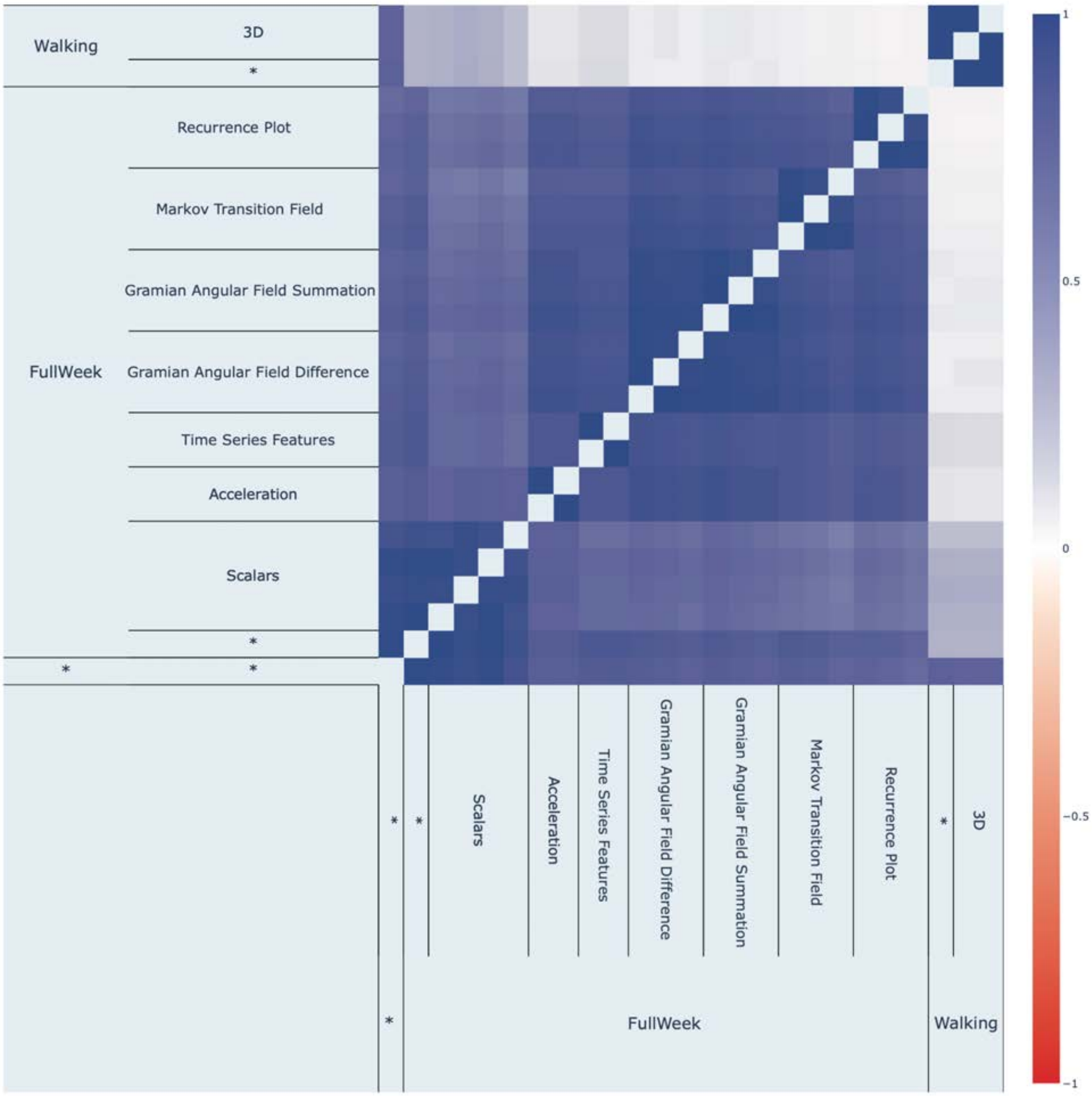

**Figure S4: Correlation between the different definitions of accelerated aging**

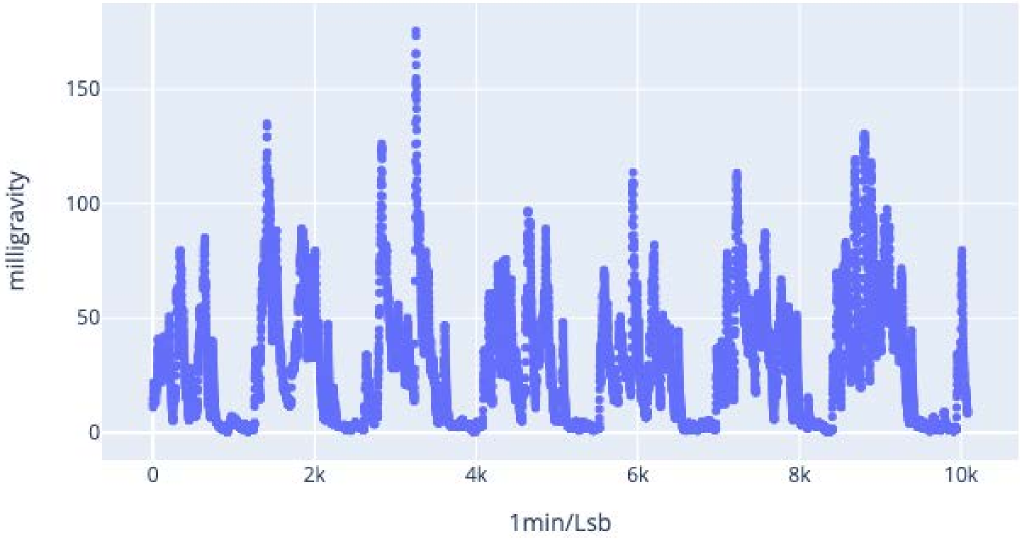

**Figure S5**: **Sample preprocessed one-dimensional acceleration time series generated from a participant's wrist accelerometer recording**

The participant is a 65-70-year-old male.

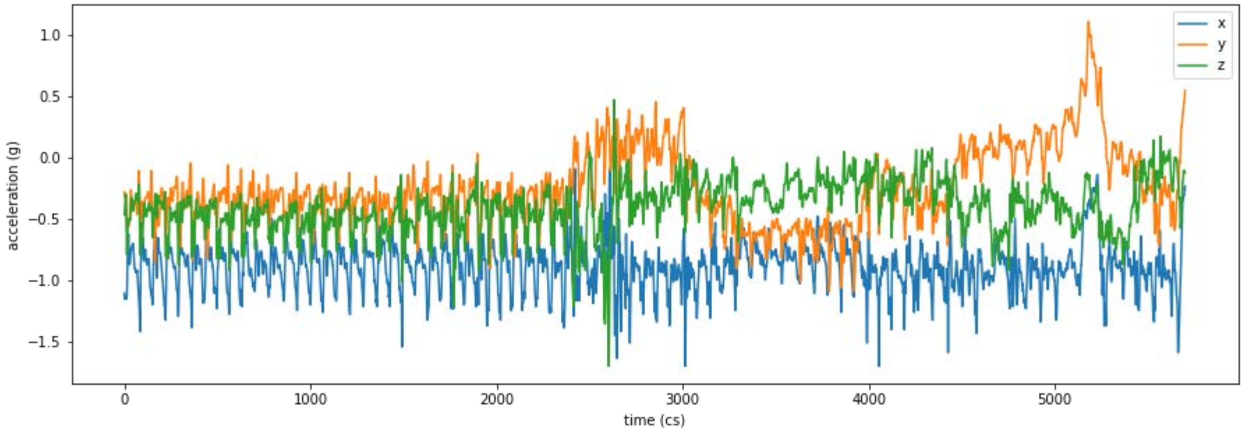

**Figure S6**: **Sample preprocessed three-dimensional acceleration time series generated from a participant's wrist accelerometer recording during their walk**

The participant is a 40-45-year-old female participant.

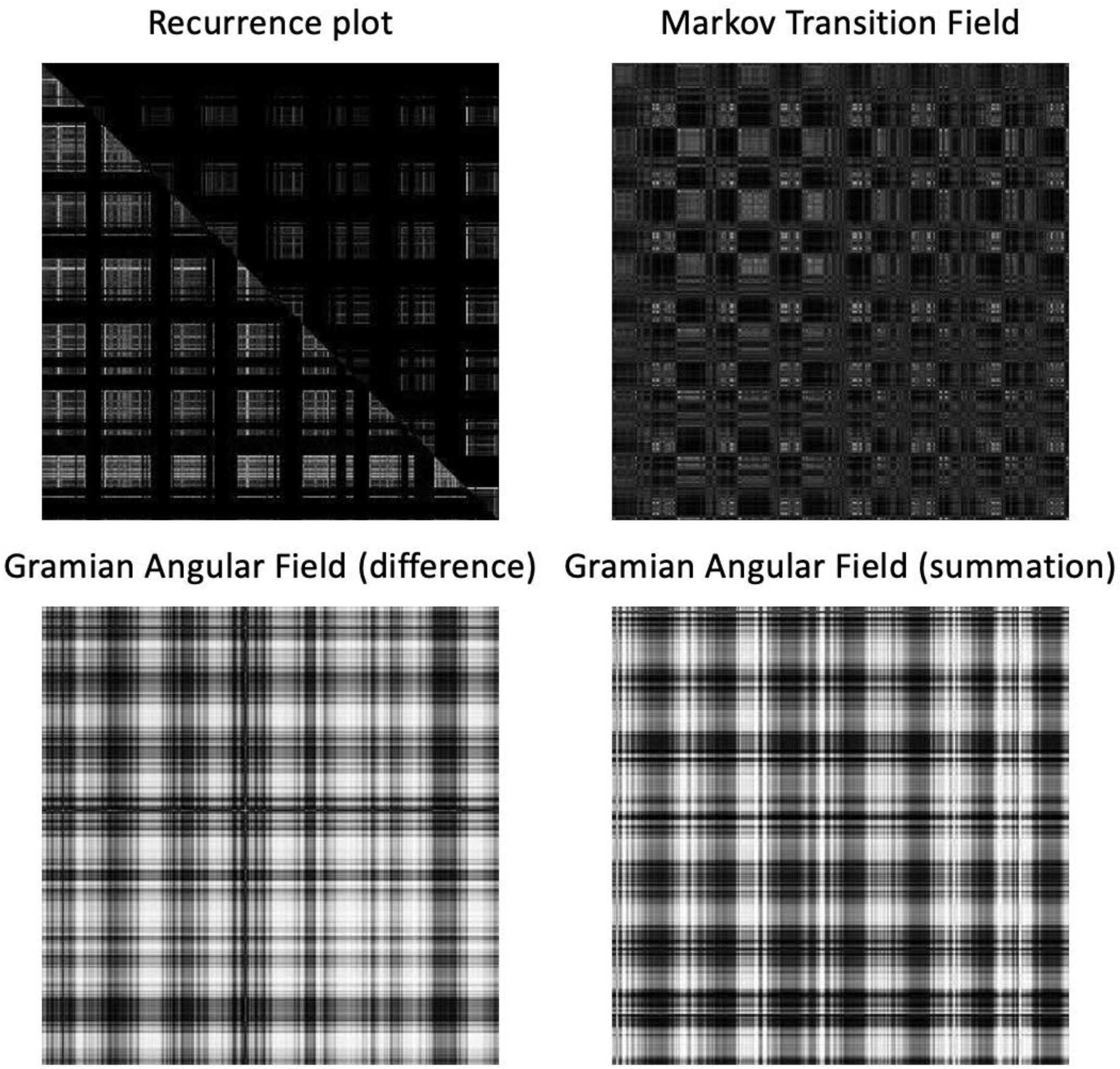

**Figure S7: Recurrence plot, Markov transition field and Gramian angular field (difference and summation) generated from a participant's wrist accelerometer recording**

The participant is a 65-70-year-old male. For the recurrence plot, the upper right triangle corresponds to the sleeping timesteps, and the lower left triangle corresponds to the remaining time steps.

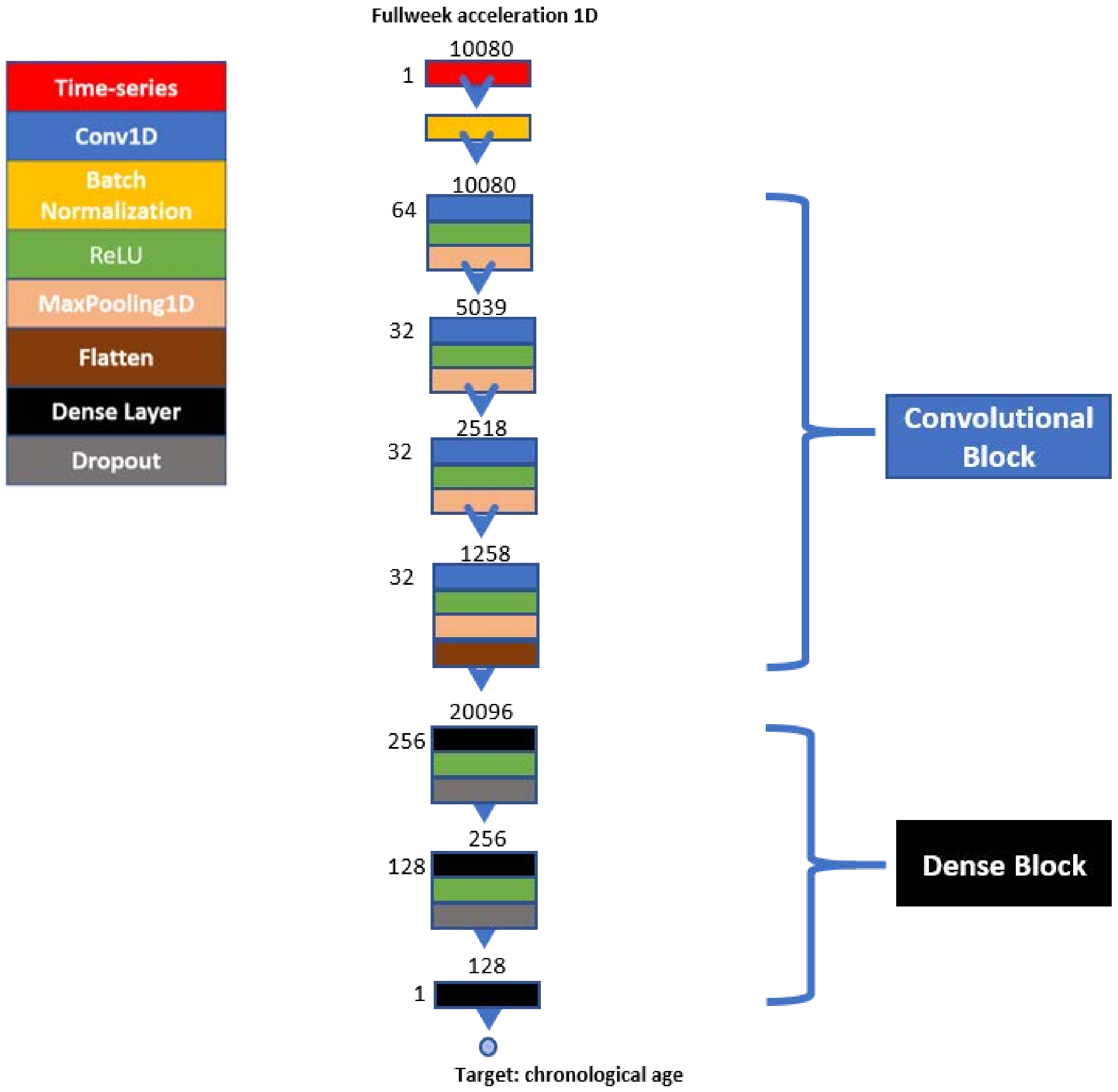

**Figure S8: Architecture of the convolutional neural network trained on the raw wrist accelerometer acceleration time series - Summary figure**

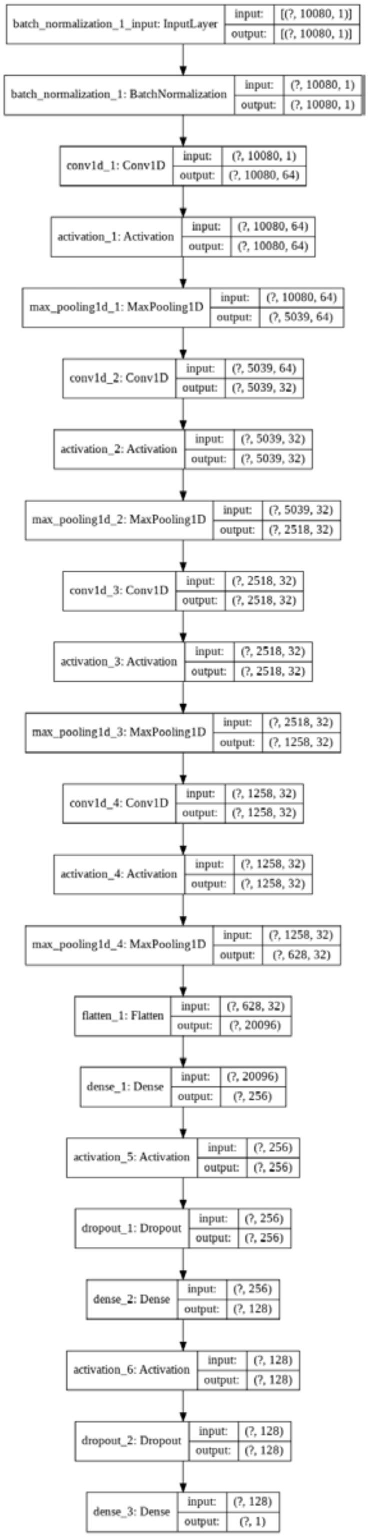

**Figure S9: Architecture of the convolutional neural network trained on the raw wrist accelerometer acceleration time series - Detailed figure**

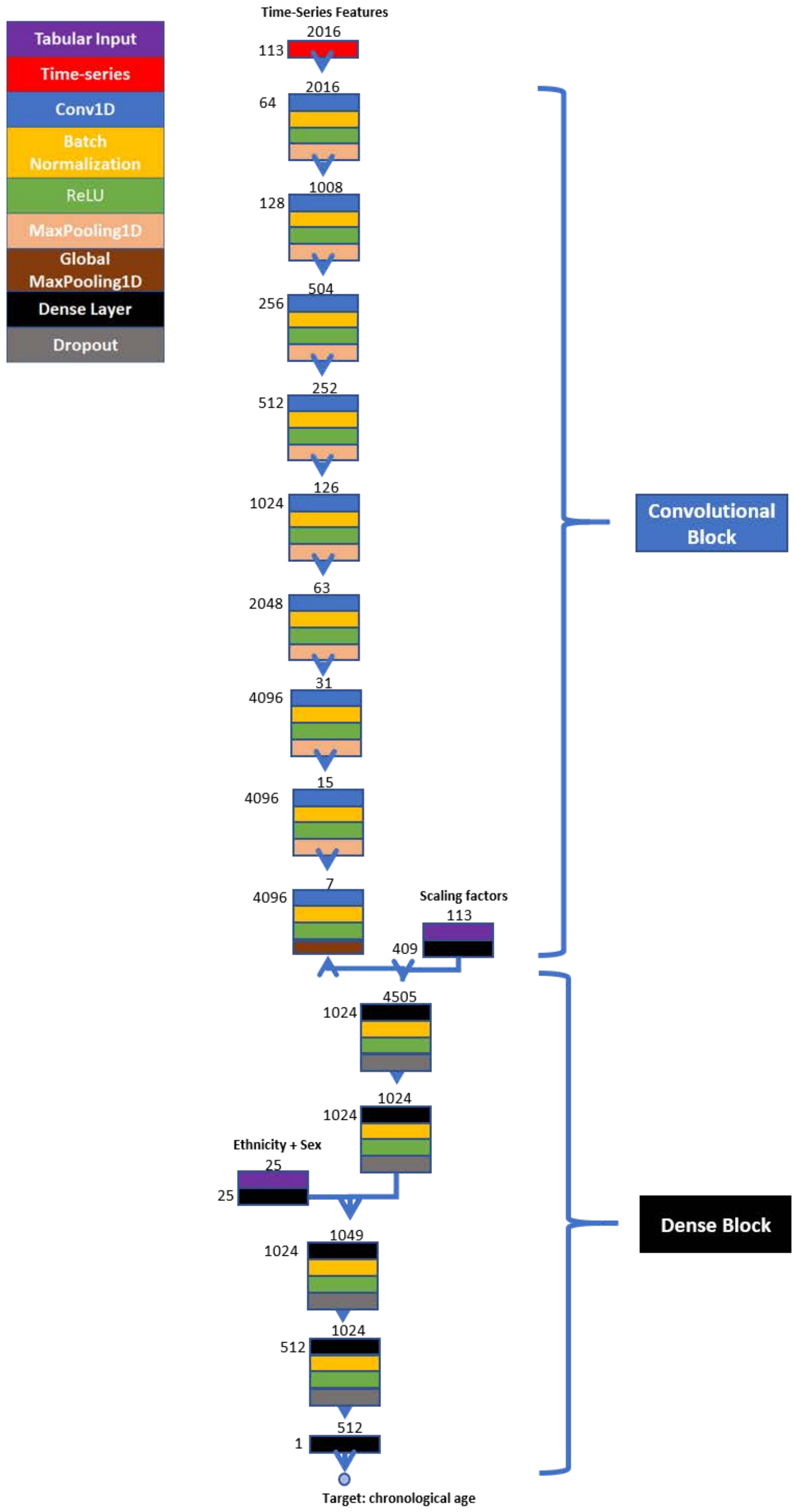

**Figure S10: Architecture of the convolutional neural network trained on the wrist accelerometer features time series - Summary figure**

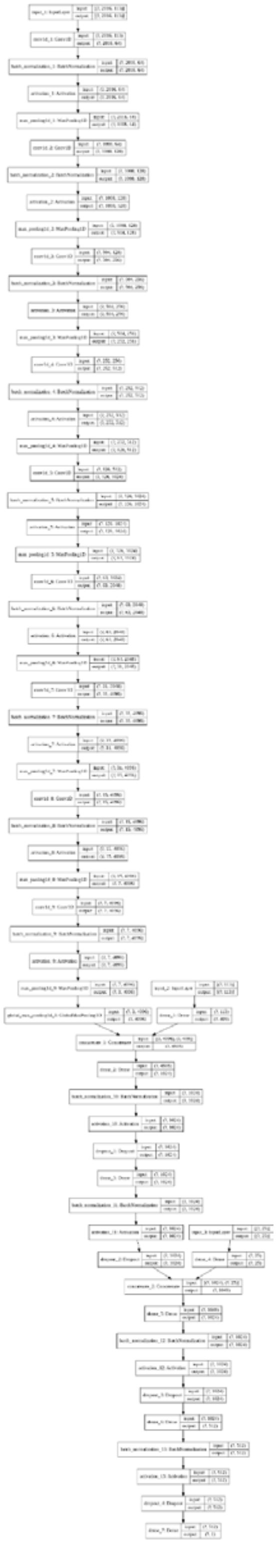

**Figure S11: Architecture of the convolutional neural network trained on the wrist accelerometer features time series - Detailed figure**

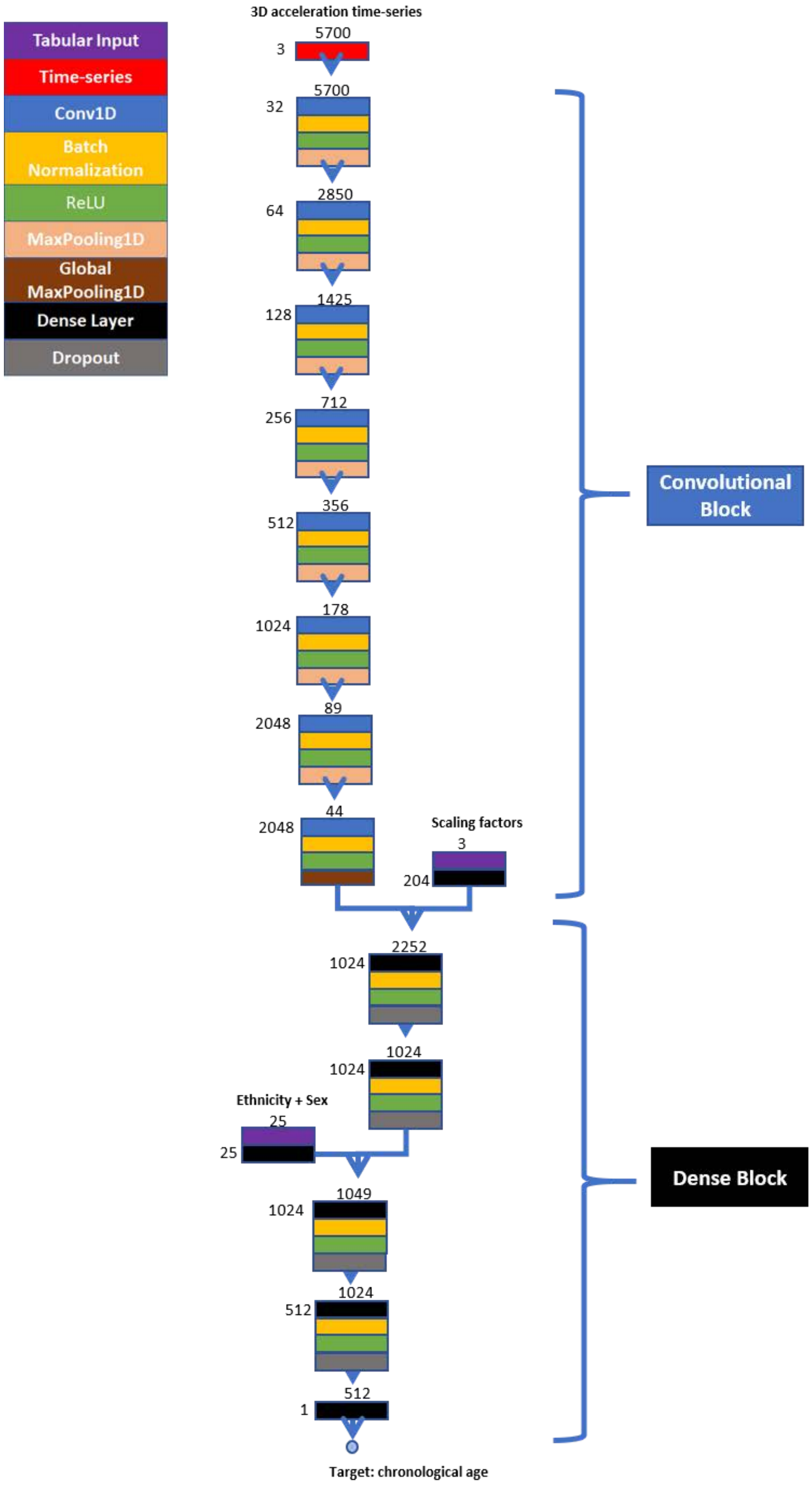

**Figure S12: Architecture of the convolutional neural network trained on the wrist accelerometer 3D raw acceleration time series - Summary figure**

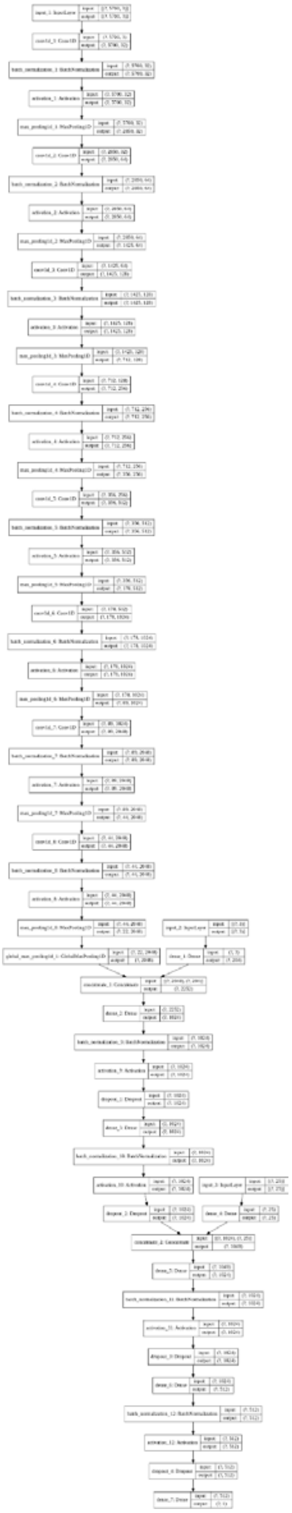

**Figure S13: Architecture of the convolutional neural network trained on the wrist accelerometer 3D raw acceleration time series - Detailed figure**

### Supplementary Tables

##### Table S1: Scalar features generated from the wrist accelerometer data

See supplementary data

##### Table S2: Feature importances for the models built on scalar predictors

See supplementary data

##### Table S3: List of biomarkers by subcategories for the Biomarkers Wide Association Study

See supplementary data

##### Table S4: Biomarkers associated with accelerated aging

| Biomarker category | Top three associations |
|---|---|
| BloodPressure (100.0%) | Pulse rate (.082); Systolic blood pressure (.041); Diastolic blood pressure (.024) |
| AnthropometryBodySize (62.5%) | Waist circumference (.114); Hip circumference (.094); Weight (.092) |
| ArterialStiffness (50.0%) | Pulse rate (.078); Position of pulse wave notch (.063); Position of the shoulder on the pulse waveform (.046) |
| CognitiveTrailMaking (50.0%) | Duration to complete numeric path (trail #1) (.049); Duration to complete alphanumeric path (trail #2) (.045) |
| HearingTest (39.7%) | Speech-reception-threshold (SRT) estimate (left) (.042); Mean signal-to-noise ratio (SNR), (left).15 (.042); Mean signal-to-noise ratio (SNR), (left).14 (.042) |
| BloodBiochemistry (35.7%) | Cystatin C (.071); Glycated haemoglobin (HbA1c) (.068); C-reactive protein (.057) |
| BloodCount (35.5%) | Neutrophill count (.064); High light scatter reticulocyte percentage (.051); High light scatter reticulocyte count (.050) |
| ECGAtRest (33.3%) | Ventricular rate (.082) |
| EyeAcuity (25.0%) | Glasses worn/required (right).wearing (.032); Glasses worn/required (left).wearing (.031) |
| UrineBiochemistry (25.0%) | Microalbumin in urine (.035) |
| HeartSize (25.0%) | Body surface area (.084); Average heart rate (.067) |
| CognitivePairsMatching (20.0%) | Time to complete round.Second round (.030); Time to complete round.First round (.020) |

| | |
|---|---|
| AnthropometryImpedance (20.0%) | Impedance of arm (right) (.025) |
| HeartPWA (20.0%) | Heart rate during PWA (.068); Number of beats in waveform average for PWA (.061); Cardiac output during PWA (.054) |
| PhysicalActivity (19.4%) | sleep-hourOfWeekday-7-avg_V2 (.263); sleep-hourOfWeekday-7-avg_V1 (.262); walking-hourOfWeekday-10-avg_V1 (.222) |
| BoneDensitometryOfHeel (16.7%) | Ankle spacing width (.078) |
| CognitiveReactionTime (14.3%) | Mean time to correctly identify matches (.066); Mean time to press snap-button (.065) |
| BraindMRIWeightedMeans (11.1%) | Weighted-mean L3 in tract anterior thalamic radiation (right) (.063); Weighted-mean MD in tract anterior thalamic radiation (right) (.062); Weighted-mean L1 in tract superior thalamic radiation (left) (.059) |
| CognitiveFluidIntelligence (2.7%) | FI4 : positional arithmetic.Do not know (.023) |
| HandGripStrength (0.0%) | |
| BrainGreyMatterVolumes (0.0%) | |
| BrainSubcorticalVolumes (0.0%) | |
| EyeAutorefraction (0.0%) | |
| EyeIntraocularPressure (0.0%) | |
| CognitiveMatrixPatternCompletion (0.0%) | |
| CognitiveNumericMemory (0.0%) | |
| CognitivePairedAssociativeLearning (0.0%) | |
| Spirometry (0.0%) | |
| CognitiveProspectiveMemory (0.0%) | |
| CognitiveSymbolDigitSubstitution (0.0%) | |
| CognitiveTowerRearranging (0.0%) | |
| CarotidUltrasound (0.0%) | |

#### **Table S5**: Biomarkers associated with decelerated aging

| Biomarker category | Top three associations |
|---|---|
| CognitiveSymbolDigitSubstitution (100.0%) | Number of symbol digit matches attempted (.096); Number of symbol digit matches made correctly (.088) |
| HandGripStrength (100.0%) | Hand grip strength (right) (.088); Hand grip strength (left) (.086) |
| Spirometry (100.0%) | Forced expiratory volume in 1-second (FEV1) (.074); Forced vital capacity (FVC) (.071); Peak expiratory flow (PEF) (.057) |
| BoneDensitometryOfHeel (83.3%) | Speed of sound through heel (.035); Heel bone mineral density (BMD) (.031); Heel bone mineral density (BMD) T-score (.031) |
| CognitiveNumericMemory (66.7%) | Maximum digits remembered correctly (.047); Number of rounds of numeric memory test performed (.042) |
| BrainSubcorticalVolumes (64.3%) | Volume of accumbens (left) (.061); Volume of thalamus (left) (.059); Volume of thalamus (right) (.055) |

| | |
|---|---|
| PhysicalActivity (59.2%) | MVPA-hourOfDay-18-avg_V1 (.325); MVPA-hourOfDay-19-avg_V1 (.310); MVPA-hourOfWeekday-18-avg_V1 (.310) |
| BloodBiochemistry (28.6%) | HDL cholesterol (.071); Apolipoprotein A (.065); Cholesterol (.036) |
| CognitivePairsMatching (20.0%) | Number of correct matches in round.First round (.021); Number of correct matches in round.Second round (.019) |
| EyeAcuity (12.5%) | Glasses worn/required (left).none (.029) |
| BrainGreyMatterVolumes (12.2%) | Volume of grey matter in VIIIa Cerebellum (right) (.060); Volume of grey matter in VIIIa Cerebellum (left) (.058); Volume of grey matter in Ventral Striatum (left) (.056) |
| CognitiveProspectiveMemory (7.1%) | Time to answer (.028) |
| HeartPWA (6.7%) | End systolic pressure index during PWA (.046) |
| CognitiveFluidIntelligence (5.4%) | Fluid intelligence score (.035); FI4 : positional arithmetic.6 (.029) |
| BraindMRIWeightedMeans (3.7%) | Weighted-mean FA in tract posterior thalamic radiation (left) (.053); Weighted-mean ICVF in tract superior thalamic radiation (right) (.047); Weighted-mean ICVF in tract cingulate gyrus part of cingulum (right) (.047) |
| BloodCount (3.2%) | Lymphocyte percentage (.047) |
| CognitivePairedAssociativeLearning (2.2%) | Number of word pairs correctly associated (.058) |
| HearingTest (0.8%) | Triplet correct (left).1 (.024) |
| BloodPressure (0.0%) | |
| ArterialStiffness (0.0%) | |
| AnthropometryImpedance (0.0%) | |
| UrineBiochemistry (0.0%) | |
| CarotidUltrasound (0.0%) | |
| CognitiveMatrixPatternCompletion (0.0%) | |
| CognitiveReactionTime (0.0%) | |
| CognitiveTowerRearranging (0.0%) | |
| CognitiveTrailMaking (0.0%) | |
| ECGAtRest (0.0%) | |
| EyeAutorefraction (0.0%) | |
| EyeIntraocularPressure (0.0%) | |
| HeartSize (0.0%) | |
| AnthropometryBodySize (0.0%) | |

**Table S6: List of clinical phenotypes by subcategories for the Clinical Phenotypes Wide Association Study**

Supplementary data

**Table S7**: **Clinical phenotypes associated with accelerated aging**

| **Clinical phenotype category** | **Top three associations** |
|---|---|
| ChestPain (100.0%) | Chest pain or discomfort walking normally (.045); Chest pain or discomfort (.043); Chest pain due to walking ceases when standing still (.042) |
| Breathing (100.0%) | Shortness of breath walking on level ground (.087); Wheeze or whistling in the chest in last year (.032) |
| Claudication (53.8%) | Leg pain on walking (.069); Leg pain when walking uphill or hurrying (.069); Leg pain when standing still or sitting (.068) |
| MentalHealth (52.3%) | Frequency of tiredness / lethargy in last 2 weeks (.067); Nervous feelings (.063); Illness, injury, bereavement, stress in last 2 years.Serious illness, injury or assault to yourself (.058) |
| GeneralPain (50.0%) | Hip pain for 3+ months (.049); Back pain for 3+ months (.043); General pain for 3+ months (.042) |
| GeneralHealth (50.0%) | Overall health rating (.159); Long-standing illness, disability or infirmity (.133); Falls in the last year (.046) |
| CancerScreening (25.0%) | Ever had bowel cancer screening (.024) |
| Hearing (21.4%) | Hearing aid user (.036); Tinnitus severity/nuisance (.028); Hearing difficulty/problems with background noise (.025) |
| Eyesight (4.8%) | Eye problems/disorders.Diabetes related eye disease (.051); Other eye problems (.034); Age when diabetes-related eye disease diagnosed (.029) |
| SexualFactors (0.0%) | |
| Mouth (0.0%) | |

**Table S8**: **Clinical phenotypes associated with decelerated aging**

| **Clinical phenotype category** | **Top three associations** |
|---|---|
| CancerScreening (25.0%) | Most recent bowel cancer screening (.028) |
| Mouth (14.3%) | Mouth/teeth dental problems.None of the above (.023) |
| GeneralHealth (12.5%) | Weight change compared with 1 year ago.No - weigh about the same (.023) |
| GeneralPain (5.6%) | Pain type(s) experienced in last month.None of the above (.049) |
| MentalHealth (4.5%) | Risk taking (.047); Illness, injury, bereavement, stress in last 2 years.None of the above (.018) |
| Eyesight (3.2%) | Eye problems/disorders.None of the above (.044); Age started wearing glasses or contact lenses (.035) |
| SexualFactors (0.0%) | |
| Hearing (0.0%) | |
| Claudication (0.0%) | |
| ChestPain (0.0%) | |
| Breathing (0.0%) | |

#### Table S9: List of diseases by subcategories for the Diseases Wide Association Study

See supplementary data

#### Table S10: Diseases associated with accelerated aging

| Disease category | Top three associations |
|---|---|
| medical_diagnoses_I (40.3%) | I10 Essential (primary) hypertension (.098); I25 Chronic ischaemic heart disease (.051); I48 Atrial fibrillation and flutter (.046) |
| medical_diagnoses_R (39.8%) | R07 Pain in throat and chest (.041); R11 Nausea and vomiting (.037); R10 Abdominal and pelvic pain (.036) |
| medical_diagnoses_K (34.7%) | K21 Gastro-oesophageal reflux disease (.041); K44 Diaphragmatic hernia (.037); K80 Cholelithiasis (.032) |
| medical_diagnoses_Z (29.8%) | Z92 Personal history of medical treatment (.069); Z86 Personal history of certain other diseases (.065); Z51 Other medical care (.051) |
| medical_diagnoses_M (28.2%) | M79 Other soft tissue disorders, not elsewhere classified (.047); M54 Dorsalgia (.046); M19 Other arthrosis (.044) |
| medical_diagnoses_G (25.4%) | G35 Multiple sclerosis (.048); G40 Epilepsy (.042); G20 Parkinson's disease (.032) |
| medical_diagnoses_E (15.9%) | E11 Non-insulin-dependent diabetes mellitus (.075); E78 Disorders of lipoprotein metabolism and other lipidaemias (.055); E66 Obesity (.051) |
| medical_diagnoses_J (15.9%) | J18 Pneumonia, organism unspecified (.037); J22 Unspecified acute lower respiratory infection (.037); J44 Other chronic obstructive pulmonary disease (.035) |
| medical_diagnoses_N (14.6%) | N18 Chronic renal failure (.041); N17 Acute renal failure (.039); N39 Other disorders of urinary system (.033) |
| medical_diagnoses_F (9.5%) | F32 Depressive episode (.050); F17 Mental and behavioural disorders due to use of tobacco (.043); F41 Other anxiety disorders (.039) |
| medical_diagnoses_D (8.5%) | D64 Other anaemias (.041); D50 Iron deficiency anaemia (.031); D69 Purpura and other haemorrhagic conditions (.025) |
| medical_diagnoses_L (5.9%) | L03 Cellulitis (.024); L40 Psoriasis (.021); L97 Ulcer of lower limb, not elsewhere classified (.021) |
| medical_diagnoses_H (5.7%) | H36 Retinal disorders in diseases classified elsewhere (.028); H91 Other hearing loss (.023); H53 Visual disturbances (.019) |
| medical_diagnoses_Y (4.7%) | Y83 Surgical operation and other surgical procedures as the cause of abnormal reaction of the patient, or of later complication, without mention of misadventure at the time of the procedure (.047); Y84 Other medical procedures as the cause of abnormal reaction of the patient, or of later complication, without mention of misadventure at the time of the procedure (.019); Y95 Nosocomial condition (.018) |
| medical_diagnoses_C (4.5%) | C77 Secondary and unspecified malignant neoplasm of lymph nodes (.021); C83 Diffuse non-Hodgkin's lymphoma (.019); C78 Secondary malignant neoplasm of respiratory and digestive organs (.018) |
| medical_diagnoses_T (4.3%) | T81 Complications of procedures, not elsewhere classified (.041); T82 Complications of cardiac and vascular prosthetic devices, implants and grafts (.027); T84 Complications of internal orthopaedic prosthetic devices, implants and grafts (.024) |
| medical_diagnoses_W (3.7%) | W01 Fall on same level from slipping, tripping and stumbling (.020); W19 Unspecified fall (.019); W18 Other fall on same level (.017) |
| medical_diagnoses_A (2.8%) | A09 Diarrhoea and gastro-enteritis of presumed infectious origin (.024); A41 Other septicaemia |

| | |
|---|---|
| | (.020) |
| medical_diagnoses_X (2.2%) | X61 Intentional self-poisoning by and exposure to antiepileptic, sedative-hypnotic, anti-Parkinsonism and psychotropic drugs, not elsewhere classified (.017); X44 Accidental poisoning by and exposure to other and unspecified drugs, medicaments and biological substances (.016) |
| medical_diagnoses_O (1.4%) | O99 Other maternal diseases classifiable elsewhere but complicating pregnancy, childbirth and the puerperium (.017) |
| medical_diagnoses_S (1.0%) | S72 Fracture of femur (.019) |
| medical_diagnoses_P (0.0%) | |
| medical_diagnoses_Q (0.0%) | |
| medical_diagnoses_V (0.0%) | |
| medical_diagnoses_B (0.0%) | |
| medical_diagnoses_U (0.0%) | |

**Table S11: List of family history variables by subcategories for the Family History Phenotypes Wide Association Study**

See supplementary data

**Table S12: List of environmental variables by subcategories for the Environmental Wide Association Study**

See supplementary data

**Table S13**: **Environmental variables associated with accelerated aging**

| Environmental variable category | Top three associations |
|---|---|
| Sleep (57.1%) | Nap during day (.095); Sleeplessness / insomnia (.063); Daytime dozing / sleeping (narcolepsy) (.058) |
| SunExposure (30.0%) | Facial ageing.About your age (.055); Facial ageing.Do not know (.046); Time spend outdoors in summer (.043) |
| Smoking (29.2%) | Number of cigarettes currently smoked daily (current cigarette smokers) (.054); Current tobacco smoking.Yes, on most or all days (.054); Difficulty not smoking for 1 day (.053) |
| ElectronicDevices (20.0%) | Plays computer games (.039); Usual side of head for mobile phone use.Left (.018) |
| EarlyLifeFactors (18.8%) | Comparative height size at age 10.Taller (.046); Country of birth (UK/elsewhere).England (.034); Comparative body size at age 10.Plumper (.028) |
| Diet (15.5%) | Major dietary changes in the last 5 years.Yes, because of illness (.071); Processed meat intake (.048); Spread type.Other type of spread/margarine (.045) |

| | |
|---|---|
| Alcohol (13.8%) | Alcohol intake frequency. (.074); Reason for reducing amount of alcohol drunk.Illness or ill health (.072); Reason for reducing amount of alcohol drunk.Doctor's advice (.031) |
| PhysicalActivityQuestionnaire (11.4%) | Time spent watching television (TV) (.119); Types of transport used (excluding work).Public transport (.051); Types of physical activity in last 4 weeks.None of the above (.044) |
| Medication (6.1%) | Medication for cholesterol, blood pressure or diabetes.Cholesterol lowering medication (.070); Medication for pain relief, constipation, heartburn.Aspirin (.051) |

##### **Table S14**: Environmental variables associated with decelerated aging

| Environmental variable category | Top three associations |
|---|---|
| ElectronicDevices (40.0%) | Weekly usage of mobile phone in last 3 months (.089); Length of mobile phone use (.073); Hands-free device/speakerphone use with mobile phone in last 3 month (.064) |
| PhysicalActivityQuestionnaire (37.1%) | Usual walking pace (.141); Types of physical activity in last 4 weeks.Strenuous sports (.113); Frequency of strenuous sports in last 4 weeks (.102) |
| Smoking (25.0%) | Time from waking to first cigarette (.057); Age started smoking in current smokers (.054); Current tobacco smoking.No (.045) |
| EarlyLifeFactors (21.9%) | Breastfed as a baby.Yes (.033); Comparative height size at age 10.Shorter (.032); Country of birth (UK/elsewhere).Elsewhere (.031) |
| Alcohol (17.2%) | Average weekly champagne plus white wine intake (.051); Alcohol drinker status (.043); Average weekly red wine intake (.041) |
| Sleep (14.3%) | Snoring (.020) |
| Diet (12.7%) | Coffee type.Ground coffee (include espresso, filter etc) (.044); Salad / raw vegetable intake (.035); Spread type.Butter/spreadable butter (.034) |
| Medication (12.1%) | Medication for pain relief, constipation, heartburn.Ibuprofen (e.g. Nurofen) (.108); Medication for cholesterol, blood pressure or diabetes.None of the above (.107); Medication for cholesterol, blood pressure, diabetes, or take exogenous hormones.None of the above (.062) |
| SunExposure (10.0%) | Facial ageing.Younger than you are (.084); Skin colour (.023) |

##### Table S15: List of socioeconomic variables by subcategories for the Socioeconomics Wide Association Study

See supplementary data

##### **Table S16**: Socioeconomic variables associated with accelerated aging

| Socioeconomic variable category | Top three associations |
|---|---|
| OtherSociodemographics (28.6%) | Private healthcare (.043); Attendance/disability/mobility allowance.Attendance allowance (.033) |
| SocialSupport (25.0%) | Leisure/social activities.Religious group (.084); Leisure/social activities.None of the above (.040) |

| | |
|---|---|
| Household (15.0%) | Own or rent accommodation lived in.Own outright (by you or someone in your household) (.091); Gas or solid-fuel cooking/heating.A gas fire that you use regularly in winter time (.043); Own or rent accommodation lived in.Rent - from local authority, local council, housing association (.043) |
| Education (12.5%) | Qualifications.None of the above (.025) |
| Employment (9.5%) | Current employment status.Unable to work because of sickness or disability (.246); Current employment status.None of the above (.016) |

**Table S17**: **Socioeconomic variables associated with decelerated aging**

| **Socioeconomic variable category** | **Top three associations** |
|---|---|
| Employment (38.1%) | Length of working week for main job (.238); Current employment status.In paid employment or self-employed (.210); Frequency of travelling from home to job workplace (.204) |
| SocialSupport (25.0%) | Leisure/social activities.Sports club or gym (.087); Leisure/social activities.Pub or social club (.049) |
| Household (15.0%) | Average total household income before tax (.142); Own or rent accommodation lived in.Own with a mortgage (.115); Number of vehicles in household (.063) |
| OtherSociodemographics (14.3%) | Attendance/disability/mobility allowance.None of the above (.105) |
| Education (12.5%) | Qualifications.College or University degree (.048) |

**Table S18: Physical activity scalar features outputted by UKB's software and removed from our list of predictors because non-biological**

See supplementary data

**Table S19: Comparison between our age predictors and the literature in terms of prediction performance**

| Physical activity dimension | Our model | | | | Model(s) in the literature | | | | | |
|---|---|---|---|---|---|---|---|---|---|---|
| | R-Squared (%) | RMSE (years) | Sample size | Age range (years) | R-Squared (%) | RMSE (years) | Algorithm | Sample size | Age range (years) | Authors |
| Daily life | 63.5±0.2 | 4.71±0 .01 | 77,269 | 43.5-79.0 | 56; 38.4 | 14; 15.74 | CNN; CNN-LSTM | 7,454; 4,268 | 6-84; 18-84 | Pyrkov et al.; Rahman and Adjeroh |
| Gait | 32.3±0.2 | 5.69±0 .01 | 98,215 | 43.5-79.0 | N/A; N/A; N/A; 88.82% (classification accuracy); 37.21; N/A | 3.02; 8.2; N/A; 14.73; 6.78 (all MAEs) | Feature engineering; Feature engineering; Random forest; Neural network; Support vector | 1,870; 1,728; 26; 142; 63,846 | 20-30; 2-94; NA (mean: 48.1, std: 12.7); 13-82; 2-90 | Lu and Tan; Makihara et al.; Riaz et al.; Hoffmann et al.; Li et al. |

| | | | | | | | |
|---|---|---|---|---|---|---|---|
| | | | | | | | machine |

**Table S20: Hyperparameter space for scalar features-based models Bayesian optimization**

| Algorithm | Hyperparameter | Scale | Low | High |
|---|---|---|---|---|
| Elastic net | alpha | loguniform | -10 | 0 |
| | l1_ratio | uniform | 0 | 1 |
| Gradient Boosted Machine | num_leaves | quniform | 5 | 45 |
| | min_child_samples | quniform | 100 | 500 |
| | min_child_weight | loguniform | -5 | 4 |
| | subsample | uniform | 0.2 | 0.8 |
| | colsample | uniform | 0.4 | 0.6 |
| | reg_alpha | loguniform | -2 | 2 |
| | reg_lambda | loguniform | -2 | 2 |
| | n_estimators | quniform | 150 | 450 |
| Neural network | learning_rate_init | loguniform | -5 | -1 |
| | apha | loguniform | -6 | 3 |

**Table S21: Nested Cross-Validation pipeline**

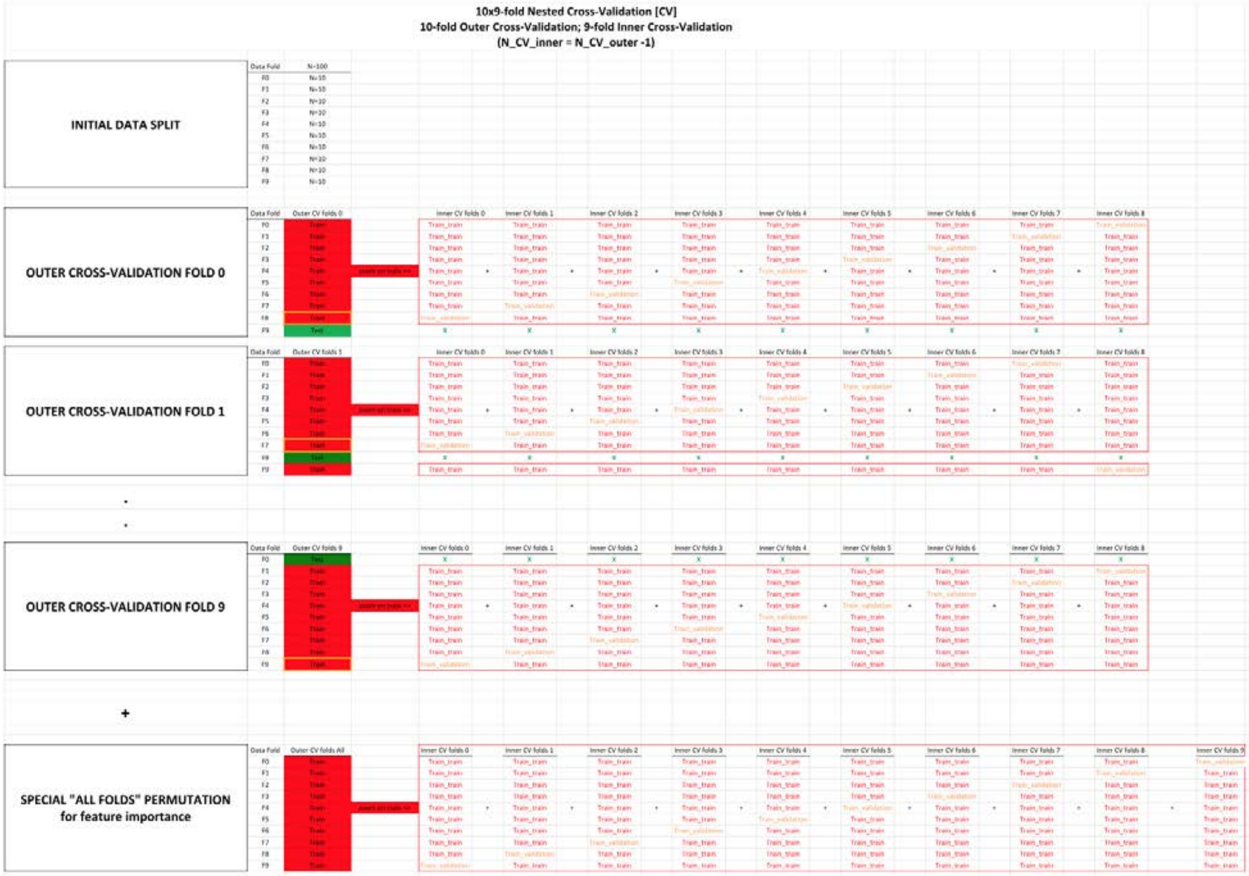

| METHODS: | |
|---|---|
| General comments | The Nested Cross-Validation is a normal double split train/val/test method, for which not just one, but the two splits are replaced with a CV instead. |
| Outer Cross-Validation | The Outer Cross-Validation is used to replace the split between train + val on one side, vs. test on the other side. Thanks to the Outer Cross-Validation, an unbiased testing prediction can be obtained for every sample.<br>Three common sources of confusion clarified:<br>**Yes**, using an Outer Cross-Validation does mean that the predictions do not all come from the same model. (For exemple for testing, the prediction for each fold comes from a different model.)<br>**No**, the Outer Cross-Validation has **"nothing"** to do with hyperparameter tuning or model selection.<br>Tuning a model only requires one Outer Cross-Validation. In contrast, it requires N_CV_folds distinct Inner Cross-Validations, one for each fold of the Outer Cross Validation. (see below) |
| Inner Cross-Validation | The Inner Cross-Validation is used to replace the split between the typical training set and validation set.<br>Thanks to the Inner Cross-Validation, hyperparameter selection can be decided based on a performance calculated on the full training set and validation set sample size, instead of the sole validation set sample size.<br>Two common sources of confusion clarified:<br>#1 - The whole purpose of performing an Inner Cross-Validation is **"hyperparameter selection"**.<br>The Inner Cross-Validation tunes the model that will then be used to generate predictions on the left out testing fold.<br>Therefore, the Inner Cross-Validation does **"not"** generate a testing prediction for more samples. That is the role of the Outer Cross-Validation.<br>One last time, the Inner Cross-Validation is **"only"** about improving the hyperparameter tuning of the model.<br>#2 - Tuning a model requires N_CV_folds distinct Inner Cross-Validations, one for each fold of the Outer Cross Validation. In contrast, it only requires one Outer Cross-Validation. (see above) |
| Special Fold 0 | Special "All folds" permutation is only here for feature importance purposes.<br>We could extract the features importance from the 10 models used to generate the predictions on the testing set, but we would have to take the average of features importances between several model.<br>Additionally each of these models only saw 90% of the data (10% was left aside for testing purposes).<br>If we only care about feature importance, we actually do not need to leave samples aside for testing. We can use 100% of the data to train a model using a 10-fold (not 9-fold this time, since the test fold can be used too!) **"Inner"** Cross-Validation to train a single model.<br>This model will never be tested, but it does not matter: its role is to provide the best estimate for the feature importance, which it does well.<br>Two common sources of confusion clarified:<br>#1 - The Nested Cross-Validation pipeline and the Special "All Folds" permutation pipeline are **"not"** intertwined and serve different purposes.<br>The purpose of the Nested Cross-Validation pipeline is to generate accurate and a unbiased testing prediction for every sample, whereas the purpose of the Special "All Folds" pipeline is to generate feature importance.<br>#2 - **Yes**, it does mean that the feature importance reported do not correspond to any of the 10 models (trained in the 10 Outer Cross-Validation folds) that generated the final testing predictions.<br>The feature importance comes from a model that was never used to generate predictions. |

| RESULTS: | **Example**: cells corresponding to the values used for each prediction set | **Explanation**: |
|---|---|---|
| Training | "E17+G17+I17+K17+M17+O17+Q17+S17+<br>E29+G29+I29+K29+M29+O29+Q29+U29+..." | To obtain the training prediction for the Data Fold F1, take the mean of every training prediction available on this data fold (10*8=80 of them). |
| Validation | "E35" | To obtain the validation prediction for the Data Fold F1, take the prediction outputted by the model trained on the remaining 8 data folds with the best hyperparameters values obtained for this Outer Cross-Validation fold.<br>These validation predictions can then be used to tune the ensemble models. /!\ Warning /!\: This can only be properly done if data folds split used for the Outer Cross-Validation and the Inner Cross-Validation is the same. |
| Testing | "C26" | No extra step is needed for the testing prediction for the Data Fold 1. Simply take the only testing prediction available for this fold. |
| Feature importance: | N/A | Extract the features importance from the special "outer fold" "All".<br>If want to compute the standard deviation for the feature importance as well, we can leverage the feature importance computed on each of the Outer Cross-Validation folds and take the square root of their variance. |

SUPPLEMENTARY:

**Comparison between Nested Cross-Validation and other validation methods:**

**#1**-Single split train/test.
Split train/test works when there are no hyperparameters to tune. If one uses a simple train/test split and tune the hyperparameters on the testing set, some overfitting is to be expected.

**#2**-Double split train/validation/test.
Because of the above, a train/validation/test double split is commonly used when hyperparameters tuning is required.
The training is performed on the training set, the tuning of the hyperparameters on the validation set, and the evaluation of the unbiased model performanc on the untouched test dataset.

**#3**-Cross-Validation(train/val) and split (train+val)/test.
With the pipeline #2 (double split train/validation/test), the tuning of the hyperparameters is not leveraging the dataset as much as with a Cross-Validation, as only one data fold is used to tune the hyperparameters.
Therefore machine learning practitioners usually replace the "inner split" (the training/validation split), by a Cross-Validation instead, but they keep the "outer split" (between the training+validation dataset used for their Cross-Validation and the testing set).

**#4**-Split train/val and Cross-Validation((train+val)/test).
Pipeline #3 only generate an unbiased testing prediction for a fraction of the dataset (the testing set).
To generate an unbiased prediction on every sample of the dataset, pipeline #4 can be used.
However, because there is a simple split between the training set and the validation set (one for each Cross-Validation fold), that means that the hypperparameter tuning only leverages a fraction of the dataset.
Pipeline #4 is preferred when generating an unbiased testing prediction for every sample is needed but the models are too time consuming to train to be able to afford a Nested Cross-Validation (#5).
Pipeline #4 is therefore the pipeline we used to predict chronological age using medical images or videos, for example.

**#5**-Nested Cross-Validation.
To fully leverage the whole dataset, a Nested Cross-Validation replaces both the outer split (train+val/test) and the inner splits (train/val) by a Cross-Validation, hence the nesting.
It requires the training of a large number of models, and it is therefore usually reserved for models that can be trained relatively quickly.
In our case, we used it to train our scalar features-based models (elastic nets, light gradient boosted machines, and shallow fully connected neural networks).

**Table S22: Hyperparameters tuning upstream of the cross-validation for images-based models**

See supplementary data

**Table S23: Outer Cross-Validation with inner split pipeline**

The values displayed are validation RMSE values. Lower values are associated with better hyperparameter tuning. When two values are displayed (value1/value2), the second value corresponds to the training RMSE. The architecture used was InceptionV3, with an initial learning rate of 0.001. The model was trained on the data folds 2-9, and validated on the data fold #0. The data fold #1 was set aside as the testing set and was not used.

| Data Fold | N=100 | Outer CV folds 0 | Outer CV folds 1 | Outer CV folds 2 | Outer CV folds 3 | Outer CV folds 4 | Outer CV folds 5 | Outer CV folds 6 | Outer CV folds 7 | Outer CV folds 8 | Outer CV folds 9 |
|---|---|---|---|---|---|---|---|---|---|---|---|
| F0 | N=10 | Train | Train | Train | Train | Train | Train | Train | Train | Validation | Test |
| F1 | N=10 | Train | Train | Train | Train | Train | Train | Train | Validation | Test | Train |
| F2 | N=10 | Train | Train | Train | Train | Train | Train | Validation | Test | Train | Train |
| F3 | N=10 | Train | Train | Train | Train | Train | Validation | Test | Train | Train | Train |
| F4 | N=10 | Train | Train | Train | Train | Validation | Test | Train | Train | Train | Train |
| F5 | N=10 | Train | Train | Train | Validation | Test | Train | Train | Train | Train | Train |
| F6 | N=10 | Train | Train | Validation | Test | Train | Train | Train | Train | Train | Train |
| F7 | N=10 | Train | Validation | Test | Train | Train | Train | Train | Train | Train | Train |
| F8 | N= | Validati | Test | Train | Train | Train | Train | Train | Train | Train | Train |

| | | | | | | | | | | | |
|---|---|---|---|---|---|---|---|---|---|---|---|
| | 10 | on | | | | | | | | | |
| F9 | N=<br>10 | Test | Train | Train | Train | Train | Train | Train | Train | Train | Validati<br>on |

43. Klambauer, G., Unterthiner, T., Mayr, A. & Hochreiter, S. Self-Normalizing Neural Networks.

in *Advances in Neural Information Processing Systems 30* (eds. Guyon, I. et al.) 971–980 (Curran Associates, Inc., 2017).

44. Prechelt, L. Early Stopping - But When? in *Neural Networks: Tricks of the Trade* (eds. Orr, G. B. & Müller, K.-R.) 55–69 (Springer Berlin Heidelberg, 1998).
45. Hochreiter, S. Untersuchungen zu dynamischen neuronalen Netzen. *Diploma, Technische Universität München* **91**, (1991).
46. Hochreiter, S., Bengio, Y., Frasconi, P., Schmidhuber, J. & Others. Gradient flow in recurrent nets: the difficulty of learning long-term dependencies. (2001).
47. Alqaraawi, A., Schuessler, M., Weiß, P., Costanza, E. & Berthouze, N. Evaluating saliency map explanations for convolutional neural networks: a user study. in *Proceedings of the 25th International Conference on Intelligent User Interfaces* 275–285 (Association for Computing Machinery, 2020).
48. Selvaraju, R. R. *et al.* Grad-cam: Visual explanations from deep networks via gradient-based localization. in *Proceedings of the IEEE international conference on computer vision* 618–626 (2017).
49. Zhou, B., Khosla, A., Lapedriza, A., Oliva, A. & Torralba, A. Learning deep features for discriminative localization. in *Proceedings of the IEEE conference on computer vision and pattern recognition* 2921–2929 (2016).
50. Wang, Z. & Yang, J. Diabetic Retinopathy Detection via Deep Convolutional Networks for Discriminative Localization and Visual Explanation. *arXiv [cs.CV]* (2017).
51. Duffy, B. A. *et al.* Regression activation mapping on the cortical surface using graph convolutional networks. (2019).
52. Le Goallec, A. & Patel, C. J. Age-dependent co-dependency structure of biomarkers in the general population of the United States. *Aging* **11**, 1404–1426 (2019).
